# Prevalence and predictors of risk factors for non-communicable diseases in individuals aged 10-30 years: Findings from baseline assessment of V-CaN – A hybrid type 2 implementation effectiveness trial

**DOI:** 10.64898/2026.01.26.26344892

**Authors:** Ashwini Kalantri, Mudita Joshi, Devyani Wanjari, Radhika Sharma, Anuj Mundra, Arjun Jakasania, Harshal Sathe, Sakshi Siriah, Pramod Bahulekar, Abhishek Raut, Chetna Maliye, Rameshwar J Paradkar, Satish Kumar, Subodh S Gupta, Bishan S Garg

**Author notes:** Department of Community Medicine, Dr DY Patil Medical College, Hospital and Research Centre, Pimpri, Dr DY Patil Vidyapeeth, Pune, Maharashtra, India. Department of Community Medicine, Shri Vasantrao Naik Government Medical College, Yavatmal, Maharashtra, India (Devyani Wanjari). Corresponding author: Abhishek Raut. These authors contributed equally to this work (Ashwini Kalantri, Mudita Joshi). These authors also contributed equally to this work (Devyani Wanjari, Radhika Sharma).

## Abstract

Adolescence and early adulthood are pivotal periods for the establishment of long-term health behaviors affecting the long-term prevalence of NCDs. We aimed to estimate prevalence of behavioral and biochemical modifiable risk factors for NCDs and predictors of common risk factors for non-communicable diseases in individuals aged 10-30 years. This trial was registered under the Clinical Trials Registration India number CTRI/2020/10/028700; https://ctri.nic.in/Clinicaltrials/showallp.php?mid1=47597&EncHid=&userName=V-CaN (International Registered Report Identifier (IRRID): DERR1-10.2196/42450). In this cross-sectional survey done on 12,000 respondents, they were selected by simple random sampling using Health and demographic surveillance system as a sampling frame, we are presenting the results of behavioral, anthropometric risk factors and biochemical risk factors for a subsample of 1600. We have also created a best-fit regression model for risk factors as outcome variables. The average consumption of salt and sugar per person per day was 8.7 (± 3.1) and 44.7 (± 18.2) grams. Prevalence of smoking, smokeless tobacco, alcohol consumption, adequate physical activity, obesity, moderate stress, high stress, anxiety and depression was 0.3%, 15.3%, 1.8%, 32.4%, 7.6%, 76.3%, 4.7%, 16% and 27.5% respectively. The mean (SD) levels of lipid panel components were higher in 20-30 years. Age group, education level, occupation, marital status, alcohol consumers, smokeless tobacco consumers, stress, anxiety, depression and elevated BP significantly affected the modifiable risk factors for NCDs. The risk factor for NCDs’ prevalence is high among adolescents and frequent screening through such surveys may be considered so that timely intervention can be done to reduce the prevalence of NCDs. The current policies around tobacco consumption focuses on smoking, more stringent laws should be made about the use of chewable tobacco as well. Screening of the younger generation for various NCDs as well as their risk factors can be considered under various programs like RKSK, RBSK, NP-NCD, NFHS etc. to ensure that we recognize this as a public health concern and take timely policy-based actions.

## Introduction

The burden of noncommunicable diseases (NCDs) is escalating in low- and middle-income nations like India, which are concurrently facing the challenges of improving nutrition, controlling infectious disease, and improving maternal and child health services. Over the next 25 years, there will likely be a large increase in the burden of cancer, diabetes, chronic respiratory diseases, cardiovascular diseases, and mental disorders. By 2030, NCDs are predicted to be responsible for around 75% deaths in India [1]. According to the National Family Health Survey-5 (NFHS-5), the overall prevalence of DM in females and men is 2.9% and 4.2%, respectively, whereas the prevalence of HTN in females and males is 13.5% and 21.6%, respectively [2]. Several studies have shown that early interventions targeting modifiable risk factors for NCDs are necessary for both primary and secondary prevention of NCD [3, 4]. The Comprehensive National Nutrition Survey (CNNS) data from 2016–2018 for all 29 Indian states reveals a startlingly high prevalence of high blood pressure in Indian youth: 35% of children aged 10–12 and 25% of those aged 13–19 had blood pressure in the stage 1 or 2 hypertension range, as defined by the American Academy of Paediatrics cut points from 2017. In addition, obesity, high blood sugar, and lipid abnormalities were among the major CVD risk factors that were more prevalent in young people with high blood pressure [5].

Evidence from low- and middle-income nations suggest that community-based interventions through health promotion programs can reduce the prevalence of risk factors for NCDs. Such interventions greatly lower the hazards associated with changing one’s diet and lifestyle, drinking alcohol, smoking, and other habits [4]. 9.3% of Indian adults living in rural areas have been observed to have metabolic syndrome, which is characterized by a clustering of abdominal obesity, hypertension, impaired glucose metabolism, and dyslipidemia [6]. According to a meta-analysis, the prevalence of childhood obesity in India is 8.4% and that of childhood overweight is 12.4% [7]. According to the National non-communicable disease monitoring survey the prevalence of adolescent overweight and obesity is 6.2% and 1.8% respectively. Around 25% adolescents have inadequate physical activity and approximately 3% consume tobacco daily [8]. In Wardha, 4.3% of kids are either overweight or obese, while 57% of teenage girls have dietary deficiencies [9].

Adolescence and early adulthood are pivotal periods for the establishment of long-term health behaviors like the mean age for initiation of consuming smokeless tobacco in India is 9.9 years as per Global Youth Tobacco Survey-4 [10]. Understanding and addressing these risk factors during these formative years can significantly reduce the future burden of NCDs. Therefore, comprehensive risk factor profiling in this age group of 10-30 years is not only vital for immediate health benefits but also for curbing the long-term prevalence of NCDs. Thus, we aim to estimate the behavioral (dietary habits, obesity, tobacco use, stress, alcohol consumption, physical inactivity) and biochemical (high blood pressure, deranged lipid panel and high blood sugar) modifiable risk factors for NCDs, and predictors of common risk factors in individuals aged 10-30 years.

## Methodology

This study was conducted as a part of Vitalizing Community for Health Promotion Against Modifiable Risk Factors of Noncommunicable Diseases (V-CaN) project (a hybrid type 2 implementation effectiveness trial) implemented in Wardha district, Maharashtra, India. This trial was registered under the Clinical Trials Registration India CTRI/2020/10/028700; https://ctri.nic.in/Clinicaltrials/showallp.php?mid1=47597&EncHid=&userName=V-CaN (International Registered Report Identifier (IRRID): DERR1-10.2196/42450) on October 28, 2020. Ethics approval was obtained from the institutional ethics committee, before the start of the trial vide letter MGIMS/IEC/COMMED/79/2020. During the study, recruitment was done from 28/03/2022 to 24/09/2022 and verbal consent was taken from all respondents for the survey. Additionally, consent was also taken if the participants were willing for blood investigation. Only, those who consented for blood investigation were considered in the sampling frame for further sampling of biochemical risk factors. In the present article, we present the results of behavioral, anthropometric and biochemical risk factors assessed at the baseline. We assessed a total of 12000 individuals (aged 10-30 years) selected using stratified random sampling (stratified for gender) for the behavioral risk factors. A sampling frame was drawn from the already existing database under the Human Demographic Surveillance System (HDSS) for the study area. From these 12000 respondents, those who consented for giving blood samples, a simple random sample was drawn for assessing the biochemical risk factors. The flowchart in figure 1 provides details on the sample collection whereas the details of the methodology (including the sample size calculation, variables studied and the details of the tool) used for the study have been published elsewhere [11]. Some measures to tackle potential biases include-random sampling, using calibrated instruments for measurements, average of repeated measures taken for vitals like blood pressure and interpretation of measurements using standardized cut-offs.

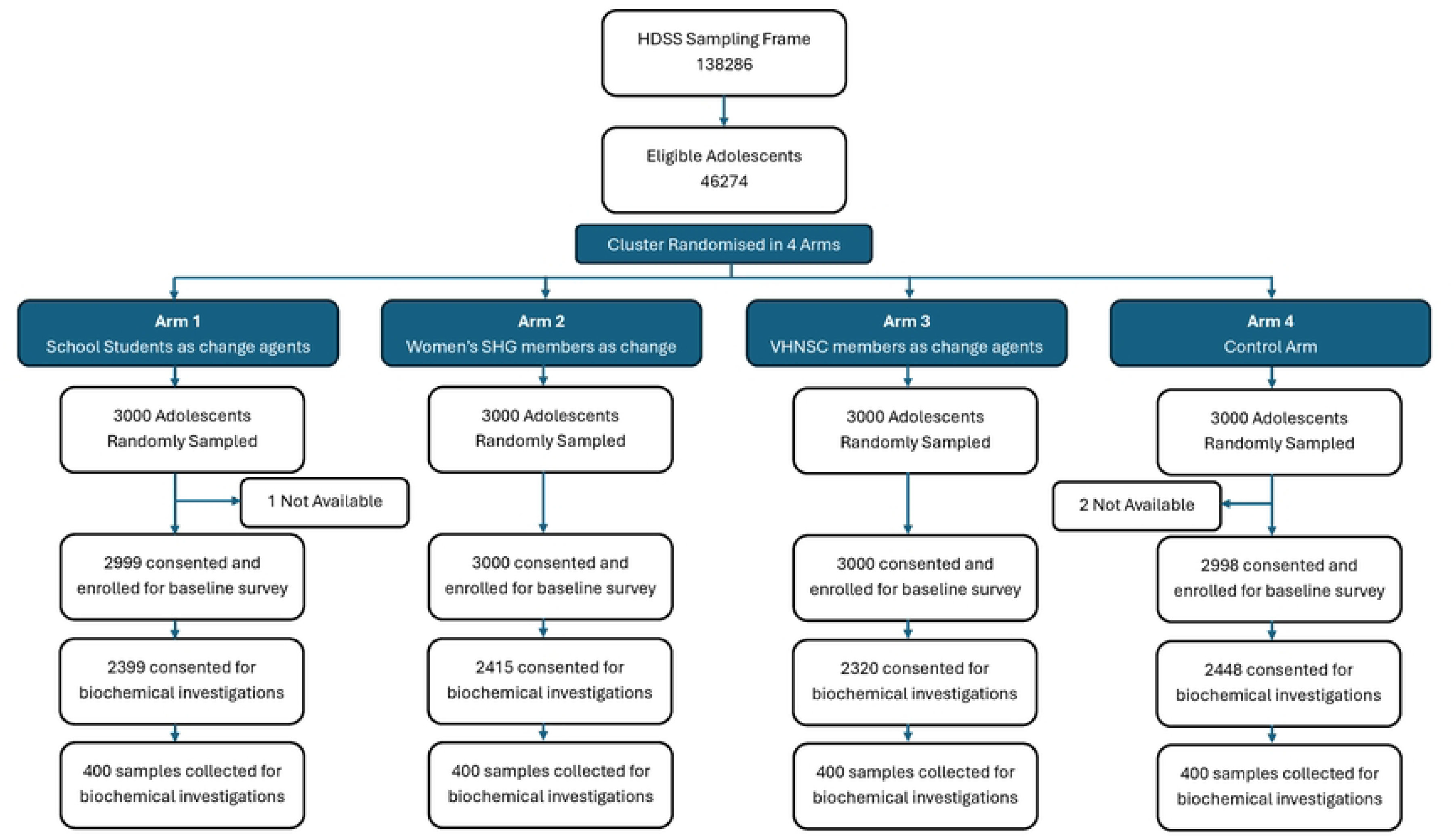

The baseline data was collected using the Survey Solutions software [12] by trained data enumerators. The data was cleaned and analyzed using R studio version 4.4.0 [13]. All of the data being reported here was collected as a part of the survey except their economic status. The information on their economic status was extracted from the database of the most recent round of the HDSS conducted in January 2023. The Wealth index was constructed using part component analysis (PCA). Categorical variables have been summarized using frequency and percentage. Continuous variables have been summarized using mean (SD) or median (IQR). Multivariate ordinal regression was performed to depict the predictors of stress. For studying the predictors of other common modifiable risk factors (smokeless tobacco consumption, physical activity, daily salt and sugar consumption) logistic regression was performed.

While analyzing and categorizing the variables under study, WHO STEPS guide was adapted to interview the respondents and we used World Health Organization (WHO) standards of recommended dietary consumption of 5 servings of fruits and vegetables per person per day [14], ≤ 5 grams of salt per person per day [15] and ≤ 50 grams of sugar per person per day [16]. For calculating the sugar and salt consumption per person per day, we divided the total salt intake with the number of family members as well as the days required to consume the packet completely. For categorizing physical activity, we have resorted to WHO reference standards suggesting a minimum of 600 met-minutes per person per day for a person to be considered as physically active as per the Global physical activity questionnaire [17]. Obesity was measured using Body mass index. Further categorization of BMI was done based on reference tables provided by WHO till 18 years of age and beyond that the Southeast Asian cut-offs were used to categorize participants based on their BMI [18]. Those respondents who were consuming any form of smokeless tobacco like kharra, betel quid, chewing tobacco, etc were reported as consumers of smokeless tobacco and those consuming alcohol in 30 days have been considered as alcohol consumers. For assessing status of hypertension, an average of three blood pressure readings were considered to draw interpretation and categorize the adolescent as normotensive, pre-hypertensive and hypertensive. This categorization was done using reference tables and cut-offs as suggested by the American academy of pediatrics [19]. To check for diabetes status, values of fasting blood sugar were assessed considering Normal level *≤ 110 mg/dl*, 110 < Pre-diabetes ≥ 126, Diabetic > 126 *mg/dl*. Stress was assessed using the perceived stress scale and was categorized based on the score as low (0-13), moderate (14-26) or high (27-40). Anxiety and depression were assessed using the patient health questionnaire-4 (PHQ-4). Those with a score of 3 or more in the first two questions were reported to have anxiety whereas those who scored 3 or more in the last two questions were reported as depressed [20]. For missing data, observations were removed from the analysis and for extreme outliers, data was imputed using the median of the variable.

We employed a forward stepwise method to sequentially add variables to the model, carefully assessing collinearity based on the Variance Inflation factor (VIF) and tolerance values at each step. For each model, bivariate analysis was done between independent variables and the outcome variables. Based on the statistical significance, we considered the variables to be added for our model. After assessing for collinearity and other model parameters, we have reported the best fitting model for the risk factors of NCDs as outcome variables-stress, smokeless tobacco consumption, physical inactivity, excess salt and sugar consumption.

## Results

The socio-demographic characteristics and risk factors have been summarized separately for the two age groups as specified earlier. Regarding the education status of the participants, 35.4% of the respondents had no or primary education whereas 12.1% had completed graduation and above. 59.5% of the respondents were students. The data on economic characteristics was available for only 9862 participants through our Health and Demographic Surveillance System (HDSS) which has been summarized as wealth quintiles. Majority of the respondents belonged to the fifth (23.6%) and fourth (23.5%) quintile of wealth index. Nearly one-fifth (20.8%) of the respondents were married at the time of data collection. Almost half (49.5%) of the respondents belonged to the OBC category followed by ST (18.7%), SC (15.4%), VJ/NT (12.3%) and Open (4.2%). The median (Inter quartile range) years of schooling was 11 (8, 12) years. (Table 1)

**Table 1:** Description of socio-demographic characteristics of the respondents.

| Characteristic (N) |  | 10-20 years<br>n(%) | 20-30 years<br>n(%) | Overall<br>n(%) |
| --- | --- | --- | --- | --- |
| **Sex (N=11,997)** | Female | 3,068 (49.5%) | 2,842 (49.0%) | 5,910 (49.3%) |
|  | Male | 3,130 (50.5%) | 2,957 (51.0%) | 6,087 (50.7%) |
| **Education**<br>**(N=11,991)** | No or primary education | 3,290 (53.1%) | 951 (16.4%) | 4,241 (35.4%) |
|  | Secondary | 1,506 (24.3%) | 1,231 (21.2%) | 2,737 (22.8%) |
|  | Higher Secondary | 1,319 (21.3%) | 2,241 (38.6%) | 3,560 (29.7%) |
|  | Graduate and above | 77 (1.2%) | 1,376 (23.7%) | 1,453 (12.1%) |
| **Occupation**<br>**(N=11,997)** | Farmer | 29 (0.5%) | 414 (7.1%) | 443 (3.7%) |
|  | Homemaker | 54 (0.9%) | 1,608 (27.7%) | 1,662 (13.9%) |
|  | Labourer | 156 (2.5%) | 1,512 (26.1%) | 1,668 (13.9%) |
|  | Service | 24 (0.4%) | 760 (13.1%) | 784 (6.5%) |
|  | Student | 5,869 (94.7%) | 1,271 (21.9%) | 7,140 (59.5%) |
|  | Unemployed | 66 (1.1%) | 234 (4.0%) | 300 (2.5%) |
| **Wealth quintiles**<br>**(N=9,862)** | Lowest | 656 (12.8%) | 495 (10.5%) | 1,151 (11.7%) |
|  | Second | 1,120 (21.8%) | 812 (17.2%) | 1,932 (19.6%) |
|  | Middle | 1,116 (21.8%) | 1,015 (21.4%) | 2,131 (21.6%) |
|  | Fourth | 1,193 (23.3%) | 1,127 (23.8%) | 2,320 (23.5%) |
|  | Highest | 1,044 (20.4%) | 1,284 (27.1%) | 2,328 (23.6%) |
| **Marital status**<br>**(N=11,997)** | Currently married | 61 (1.0%) | 2,431 (41.9%) | 2,492 (20.8%) |
|  | Divorced/ Separated | 5 (0.1%) | 42 (0.7%) | 47 (0.4%) |
|  | Never married | 6,132 (98.9%) | 3,326 (57.4%) | 9,458 (78.8%) |
| **Caste**<br>**(N=11,997)** | Open | 271 (4.4%) | 229 (3.9%) | 500 (4.2%) |
|  | OBC | 3,049 (49.2%) | 2,886 (49.8%) | 5,935 (49.5%) |
|  | SC | 973 (15.7%) | 876 (15.1%) | 1,849 (15.4%) |
|  | ST | 1,154 (18.6%) | 1,088 (18.8%) | 2,242 (18.7%) |
|  | VJ/NT/Others | 751 (12.1%) | 720 (12.4%) | 1,471 (12.3%) |
| **Years of schooling (11,997)\*** |  | 9 (7.0, 11) | 12 (10.0, 14) | 11 (8.0, 12) |
| *median (IQR) has been reported for years of schooling<br>OBC – Other Backward Class, SC- Scheduled caste, ST- Scheduled tribe, VJ– Vimukt Jati, NT- Nomadic Tribes, IQR- Inter quartile range |  |  |  |  |

We have reported the prevalence of common risk factors of non-communicable diseases such as-dietary practices, addiction, adequate physical activity, stress, measures to estimate obesity, hypertension, diabetes status (table 2a) and the biochemical measurements for a subgroup of 1600 respondents (table 2b). Among dietary practices, we have reported servings of fruits and vegetables along with daily dietary intake of salt and sugar. Based on our observations, 99.9% consumed less than 5 servings of fruits and vegetables. 95.9% of the respondents consume more than 5 grams salt per person per day which exceeds the recommended intake of 5 grams per person per day whereas 27.4% respondents consumed more sugar than the recommended limit of 50 grams.

**Table 2a:**
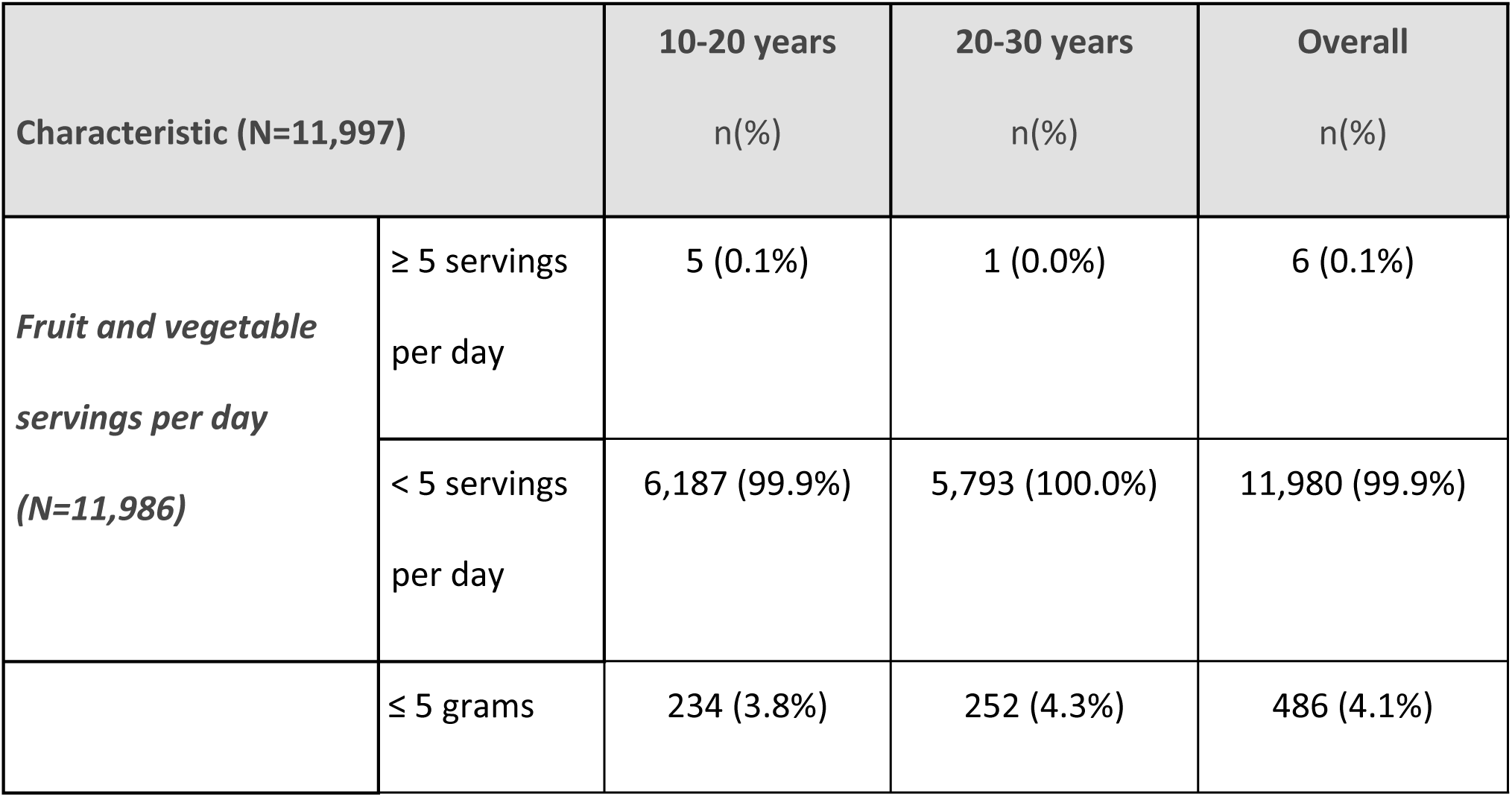
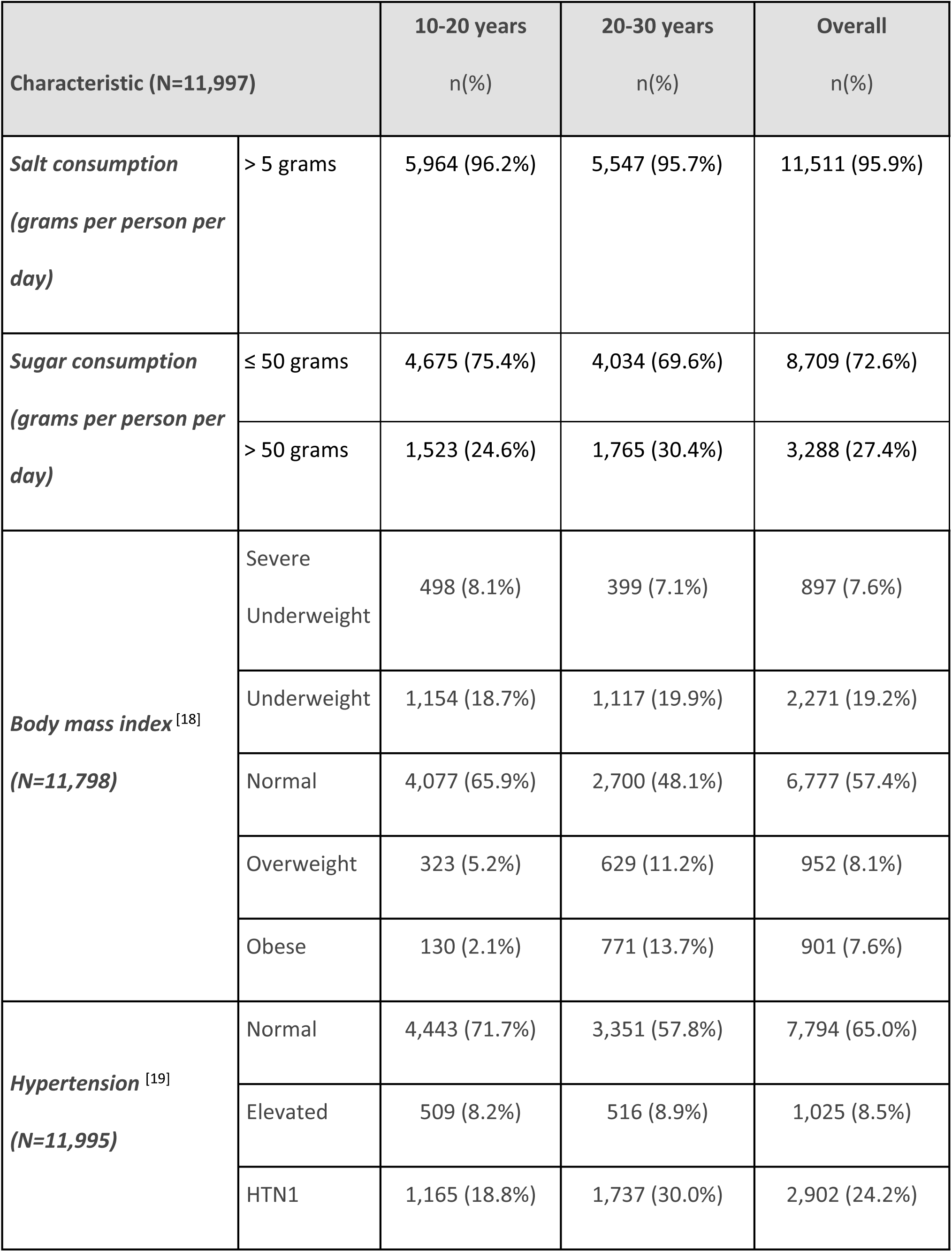
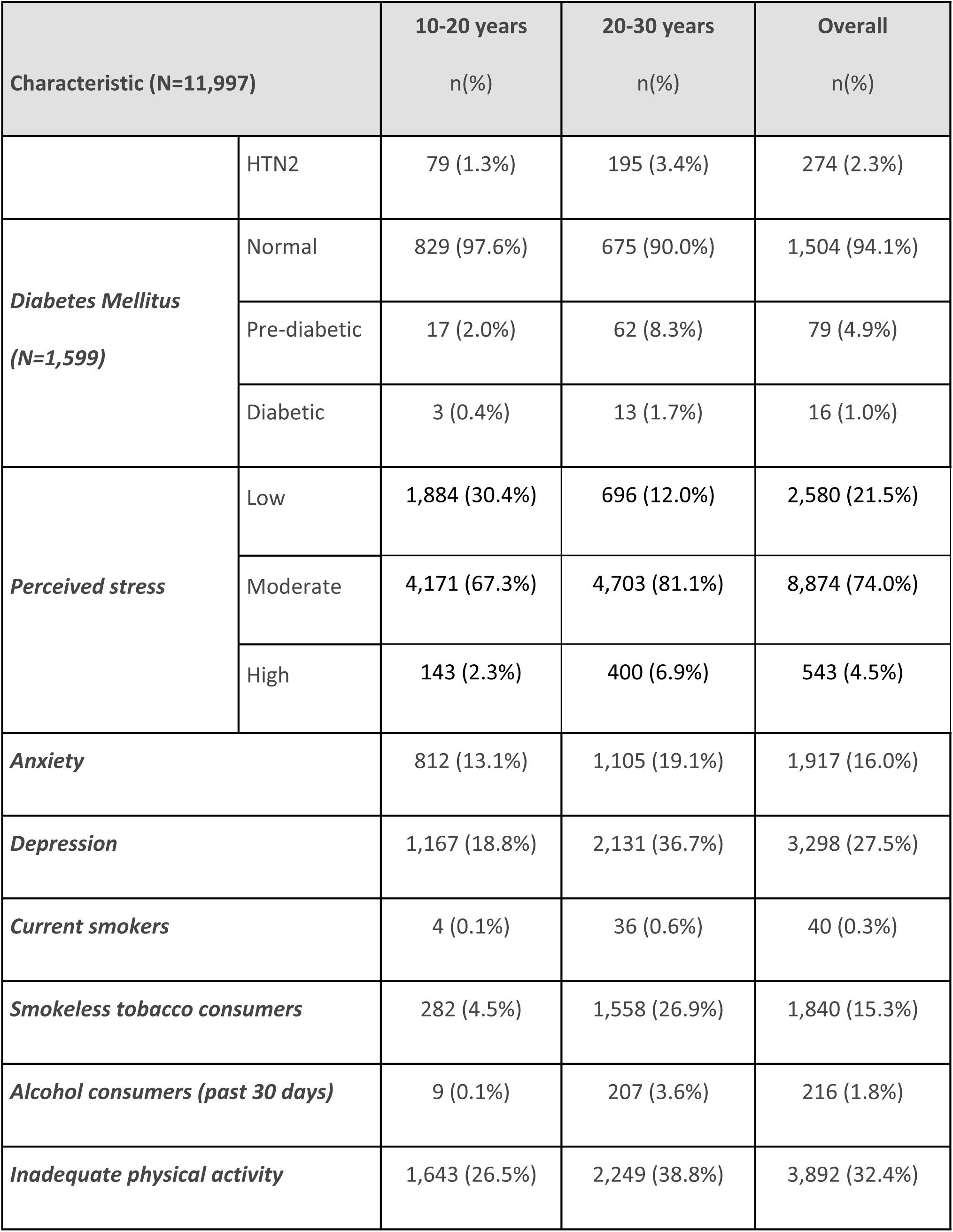
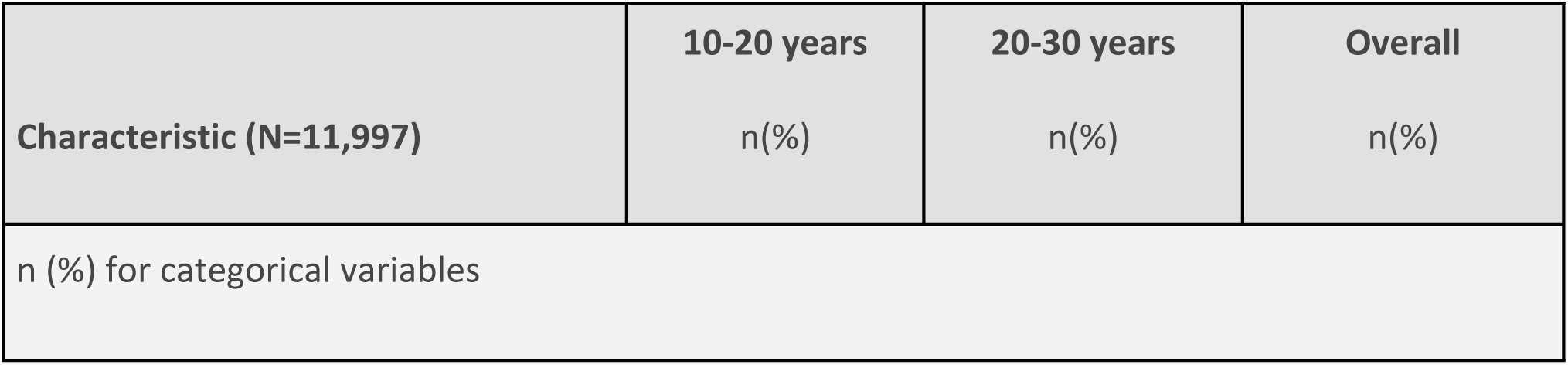
Prevalence of behavioral risk factors for NCDs.

**Table 2b:** Distribution of biochemical risk factors.

| Characteristic | 10-20 years<br>N = 849 | 20-30 years<br>N = 751 | Overall<br>N = 1600 |
| --- | --- | --- | --- |
| **Glucose<sup>1</sup> (gm/dl)** | 81.0 (12.8) | 85.2 (19.8) | 83.0 (16.6) |
| **Cholesterol<sup>1</sup> (mg/dl)** | 130.4 (26.9) | 149.2 (34.3) | 139.2 (32.0) |
| **Triglycerides (TG)<sup>2</sup> (mg/dl)** | 76.0 (59.0, 98.0) | 84.5 (62.0, 114.0) | 79.0 (60.0, 106.0) |
| **Low-density Lipoprotein (LDL)<sup>1</sup> (mg/dl)** | 71.9 (21.0) | 85.0 (26.5) | 78.1 (24.6) |
| **Very low-density Lipoprotein (VLDL)<sup>2</sup> (mg/dl)** | 15.2 (11.8, 19.6) | 16.9 (12.4, 22.8) | 15.8 (12.0, 21.2) |
| **High-density Lipoprotein (HDL)<sup>1</sup>**<br>**(mg/dl)** | 41.8 (9.7) | 44.7 (11.2) | 43.2 (10.5) |
| <sup>1</sup>Mean (SD); <sup>2</sup>Median (25%-75%). |  |  |  |

Most (57.4%) of the respondents had normal BMI, 19.2% were underweight whereas the prevalence of severe underweight, overweight and obesity was 7.6%, 8.1% and 7.6% respectively. The prevalence of overnutrition was seen to be higher in the 20-30 years age group. Nearly two-third respondents (65.0%) were normotensive, 24.2% were in HTN grade 1 and 2.3% fell in HTN grade 2. 94.1% had normal blood glucose levels, 4.9% were pre-diabetic and 1% were diabetic.

Higher levels of stress, anxiety and depression were seen among individuals aged 20-30 years. Overall 21.5% of the respondents had low stress, almost three-fourths (74.0%) had moderate stress and 4.5% had high stress levels. The overall prevalence of anxiety and depression was 16% and 27.5% respectively in the study population. Only 0.3% of our respondents reported the practice of smoking, 15.3% reported consumption of smokeless tobacco and 1.8% reported alcohol consumption in 30 days preceding the survey. Around 32.4% turned out to be physically inactive with more physically inactive participants (38.8%) from the 20-30 age group.

Table 2b suggests prevalence of biochemical risk factors. Overall, mean (SD) glucose level was 83.0 (16.6) gm/dl. The mean (SD) levels of cholesterol, LDL, and HDL were 139.2 (32.0) mg/dl, 78.1 (24.6) mg/dl and 43.2 (10.5) mg/dl respectively. We are reporting the median values of TG and VLDL here due to non-normal distribution in the study sample. The Median (IQR) value of TG and VLDL were 79.0 (60.0 - 106.0) mg/dl and 15.8 (12.0 - 21.2) mg/dl.

Table 3 shows the association between stress with the socio-demographic variables and the risk factors under study. The best-fitting model for stress as an outcome was created after adjusting for variables mentioned in the footnote of table 3. It was selected based on comparisons of the Akaike Information Criterion (AIC), Bayesian Information Criterion (BIC), and R-squared (Nagelkerke’s R² value = 0.455) values among the candidate models.

**Table 3:**
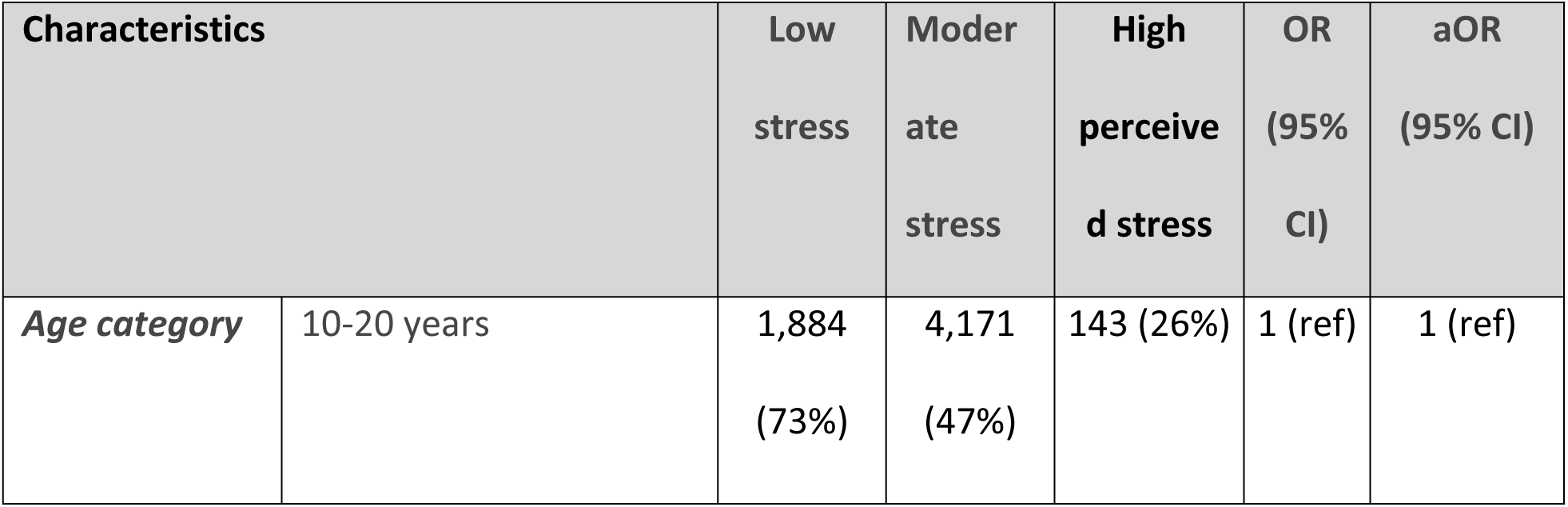
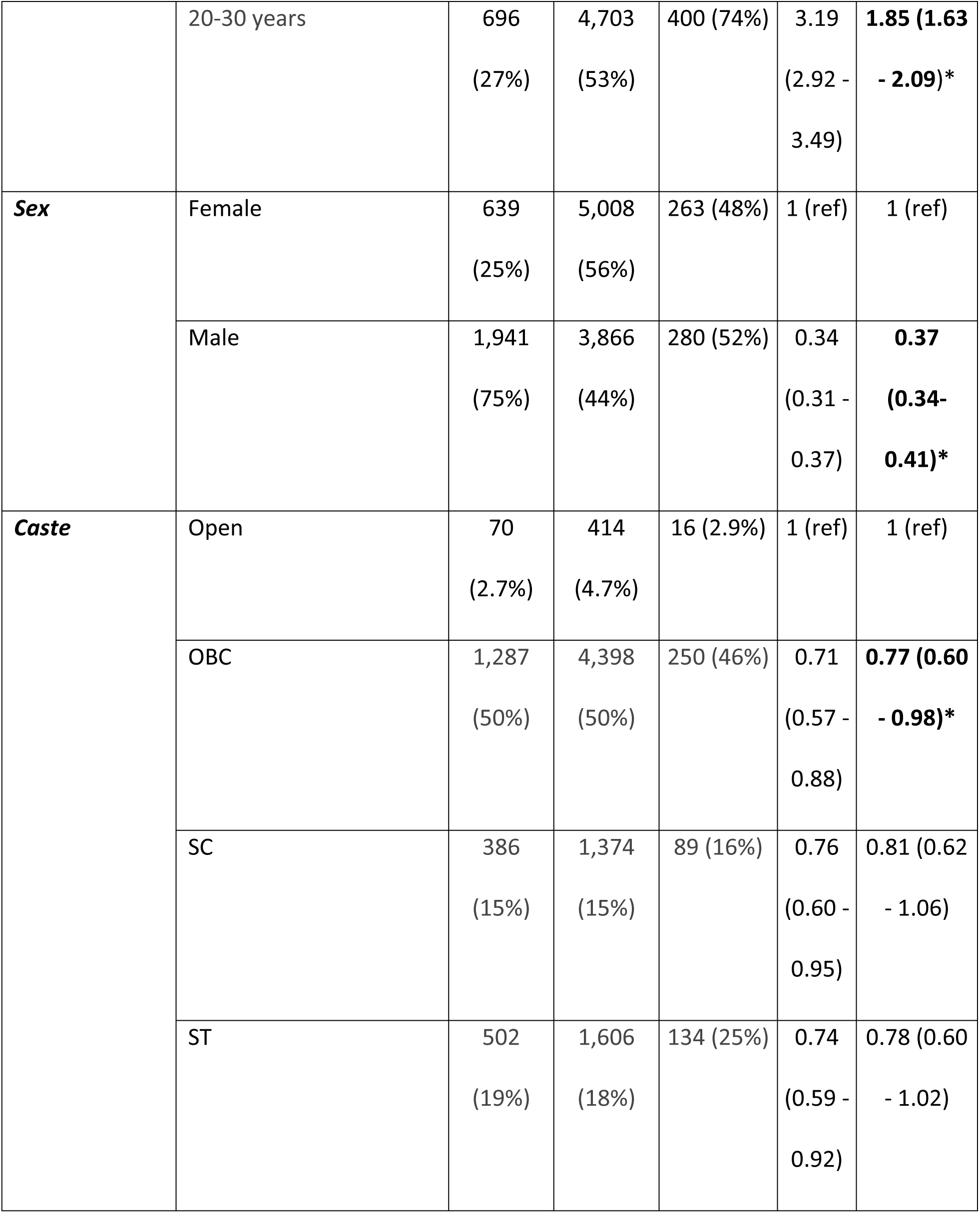
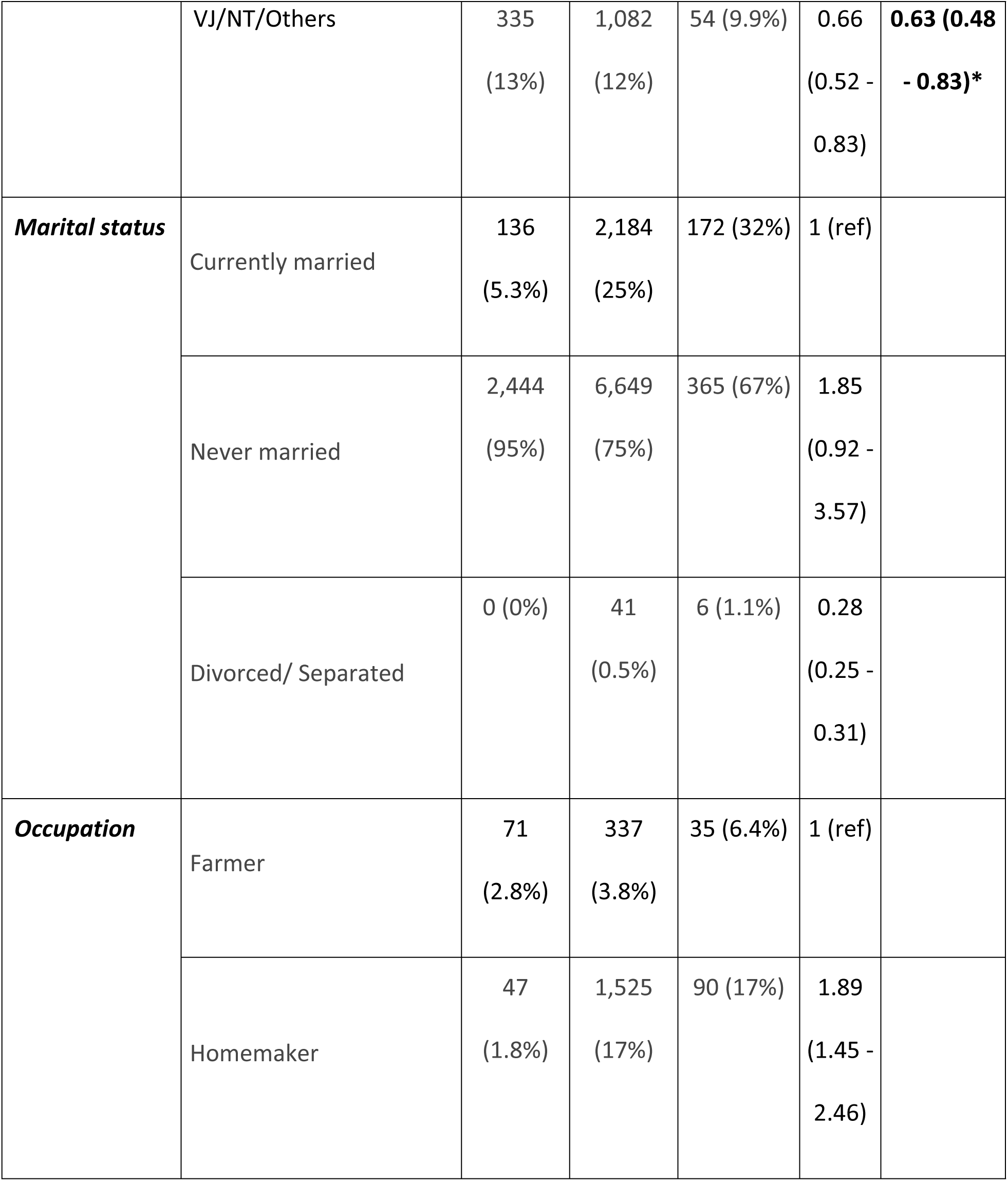
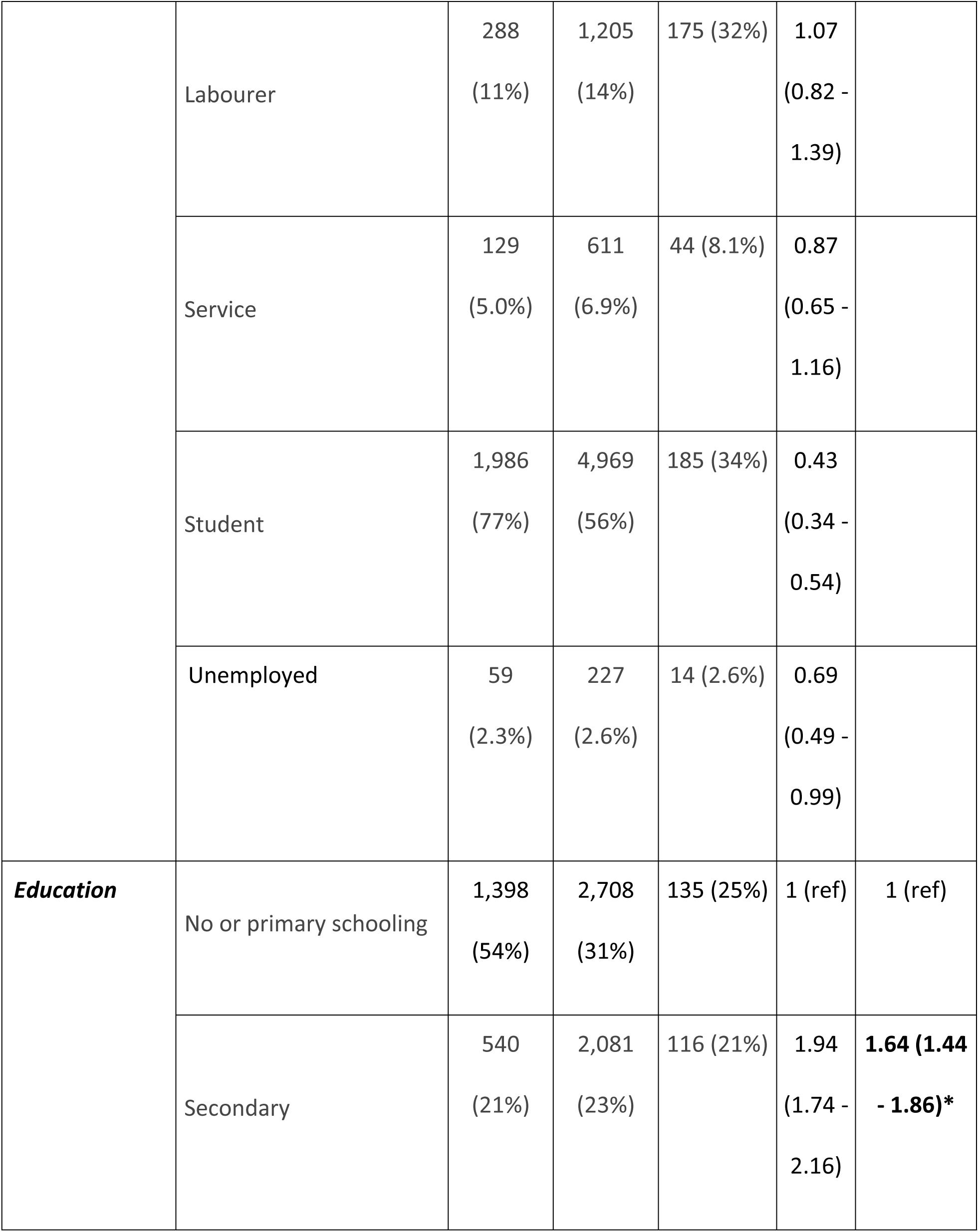
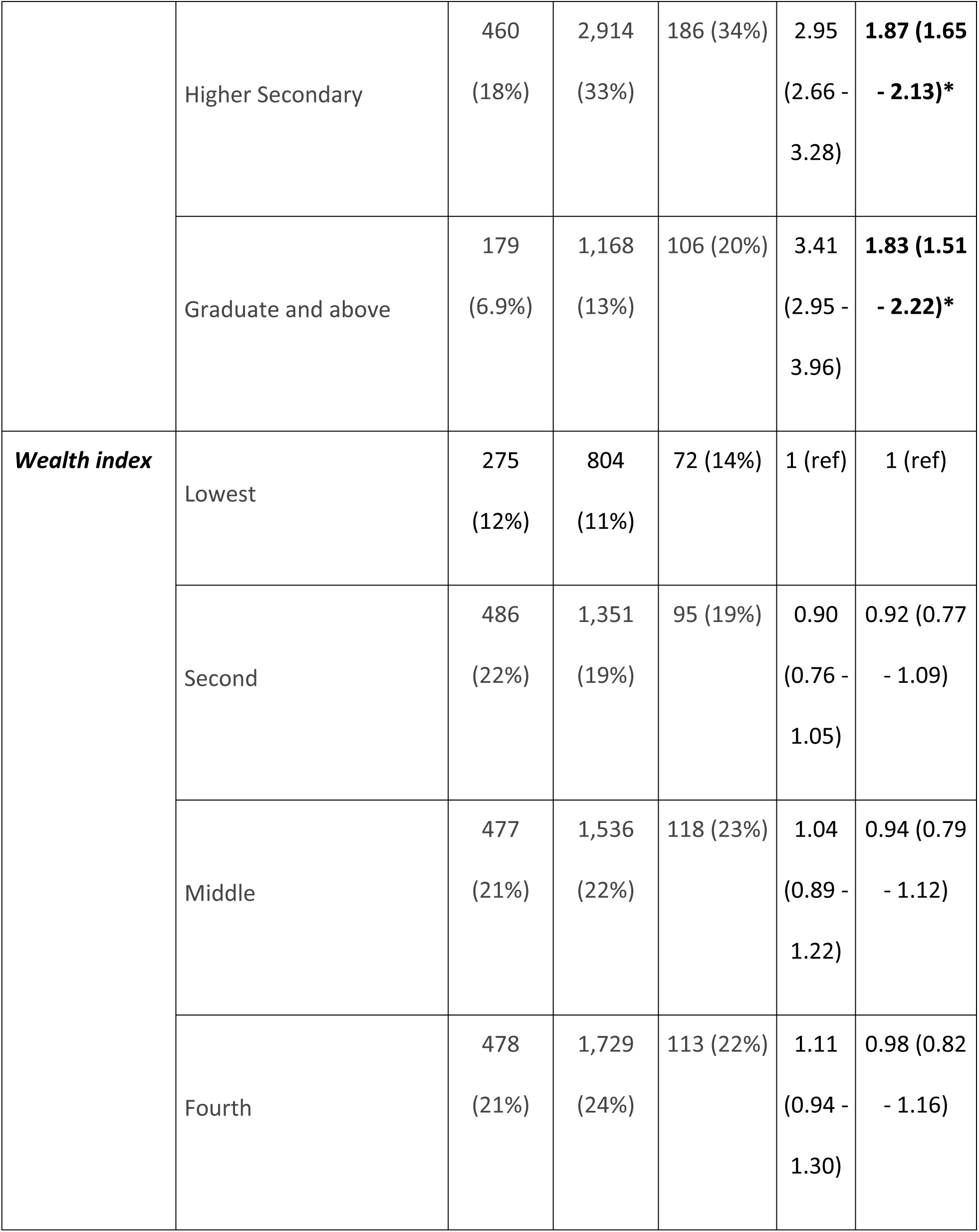
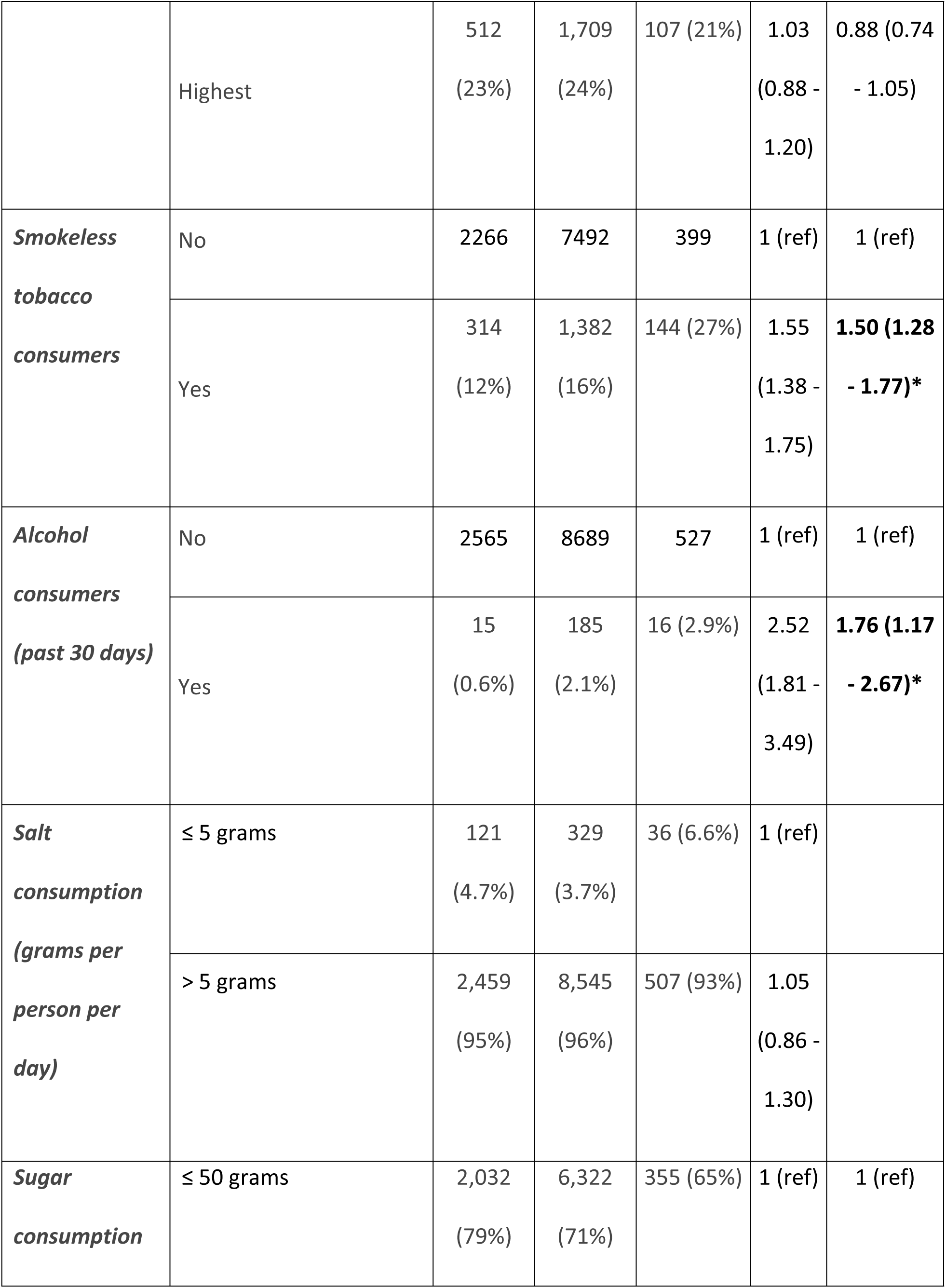
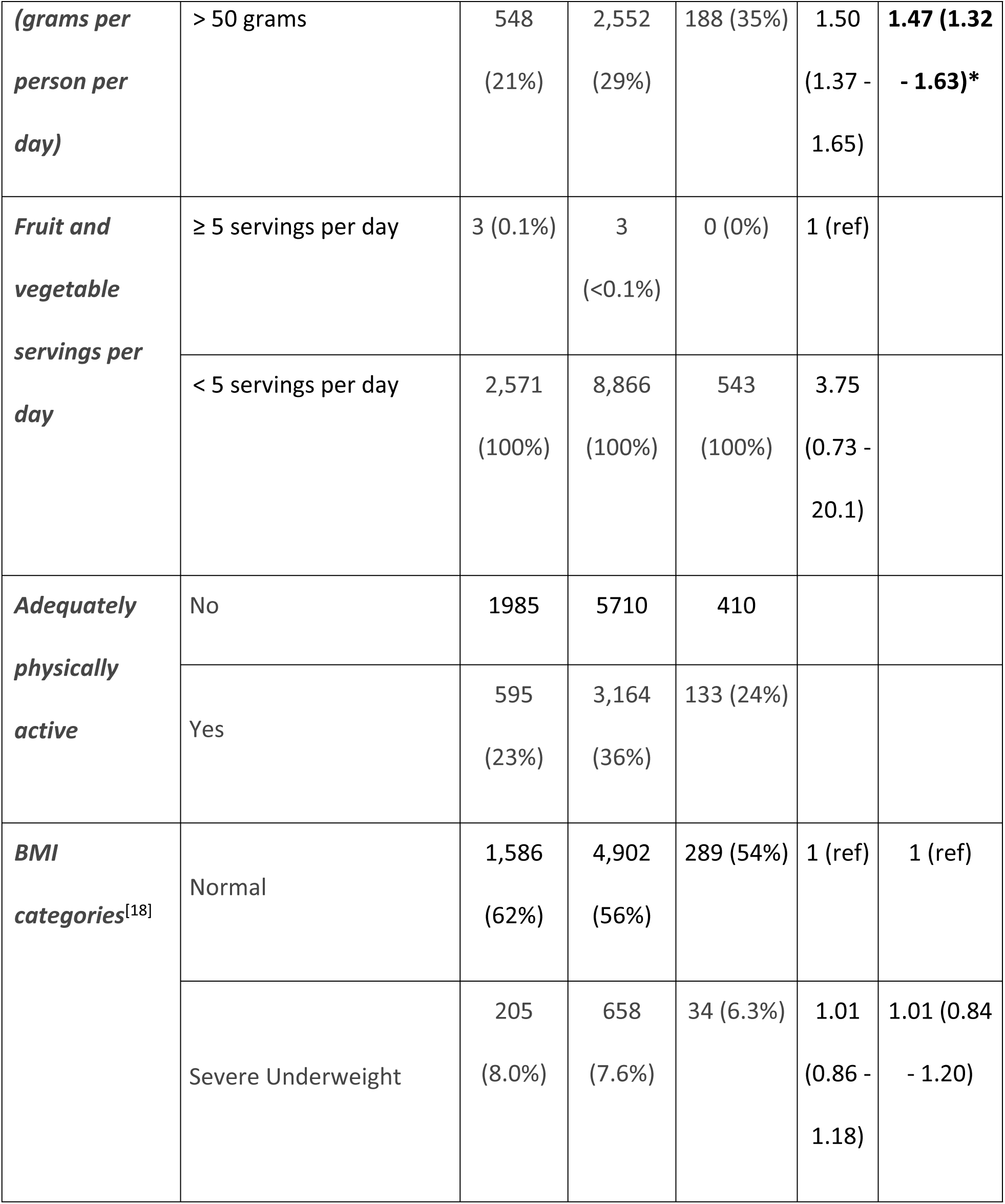
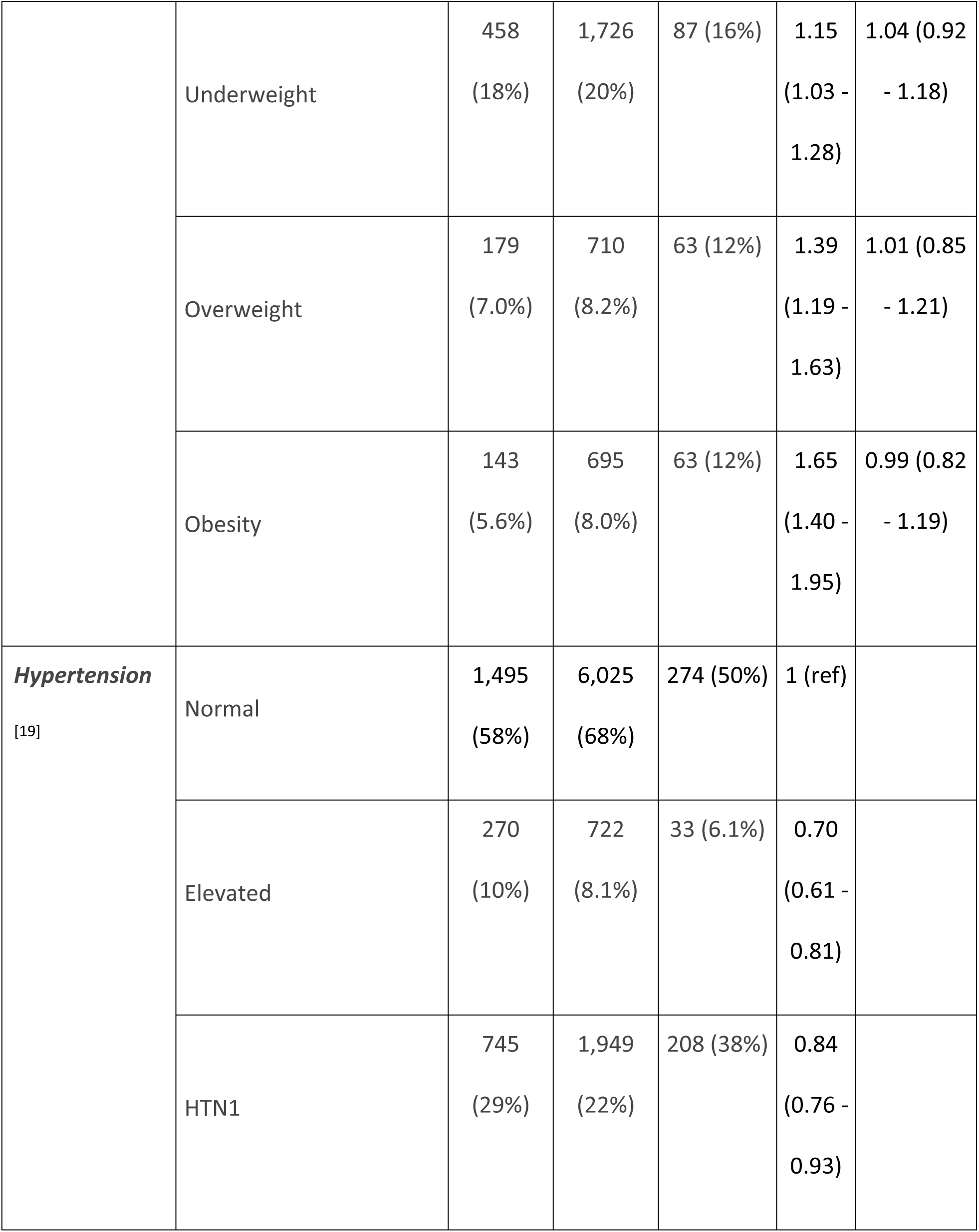
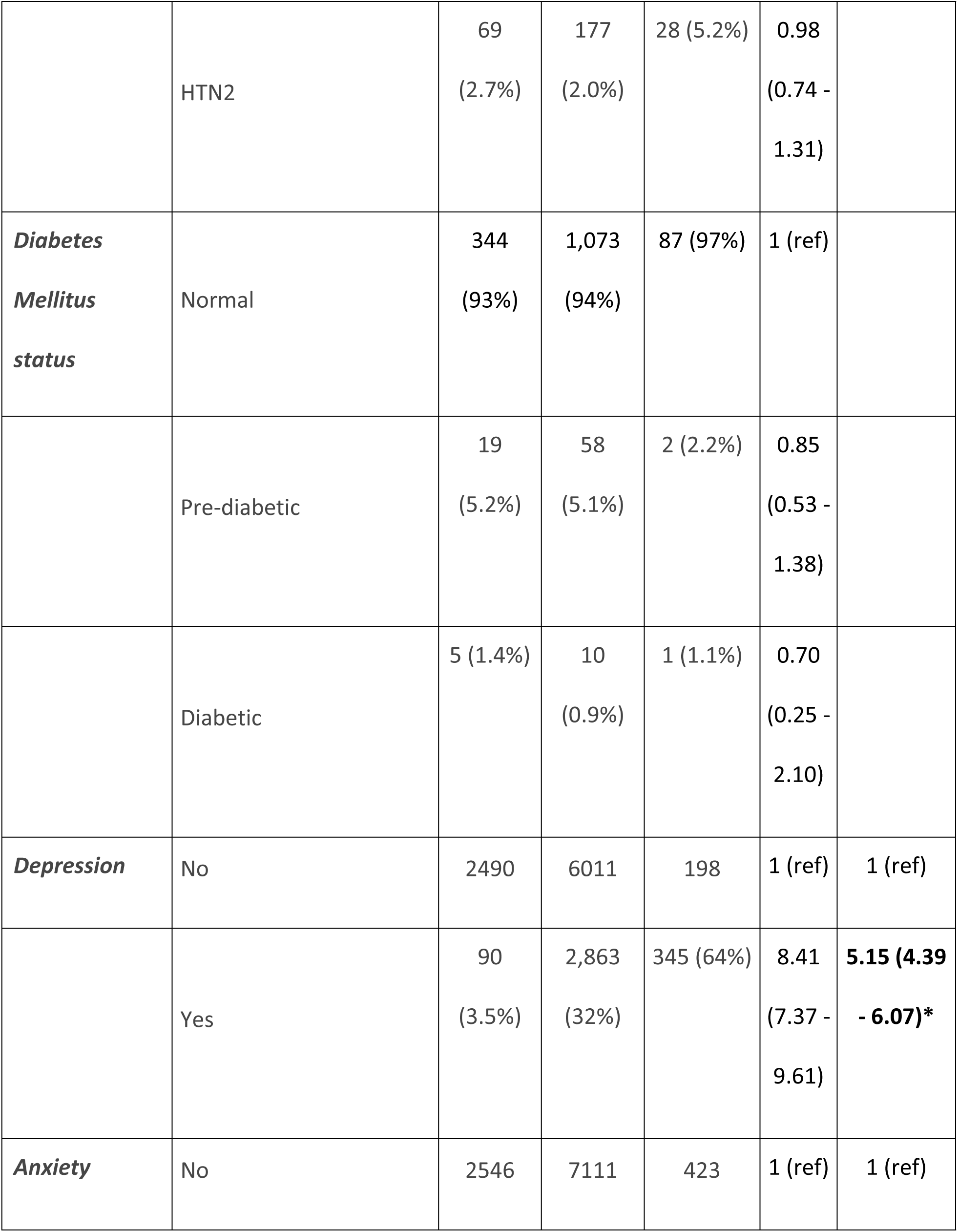
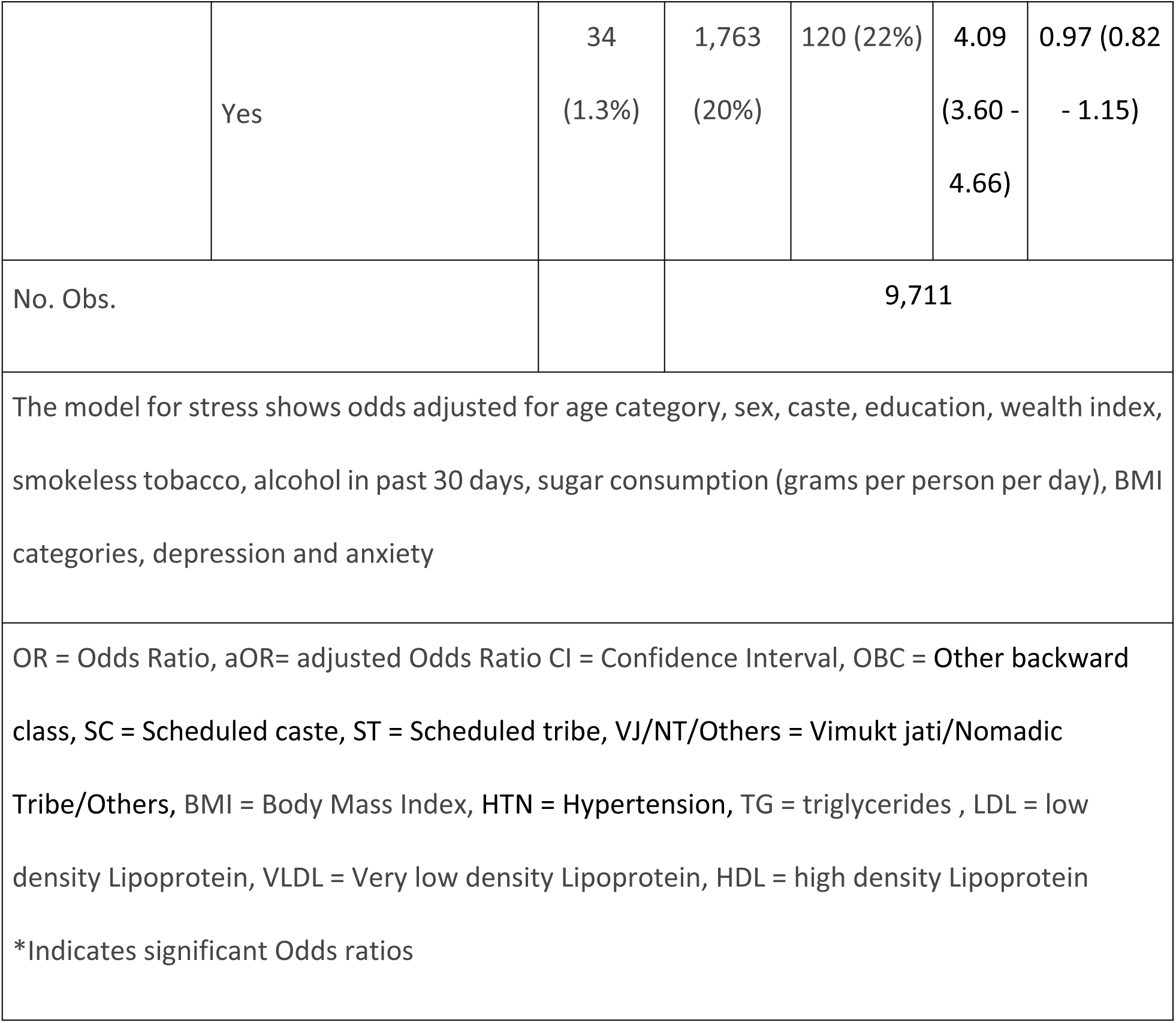
Association of stress with socio-demographic and other behavioral characteristics among 10-30 years old persons.

| Characteristics |  | Low stress | Moderate stress | High perceived stress | OR (95% CI) | aOR (95% CI) |
| --- | --- | --- | --- | --- | --- | --- |
| **Age category** | 10-20 years | 1,884 (73%) | 4,171 (47%) | 143 (26%) | 1 (ref) | 1 (ref) |
|  | 20-30 years | 696<br>(27%) | 4,703<br>(53%) | 400 (74%) | 3.19<br>(2.92 - 3.49) | **1.85 (1.63 - 2.09)\*** |
| **Sex** | Female | 639<br>(25%) | 5,008<br>(56%) | 263 (48%) | 1 (ref) | 1 (ref) |
|  | Male | 1,941<br>(75%) | 3,866<br>(44%) | 280 (52%) | 0.34<br>(0.31 - 0.37) | **0.37 (0.34-0.41)\*** |
| **Caste** | Open | 70<br>(2.7%) | 414<br>(4.7%) | 16 (2.9%) | 1 (ref) | 1 (ref) |
|  | OBC | 1,287<br>(50%) | 4,398<br>(50%) | 250 (46%) | 0.71<br>(0.57 - 0.88) | **0.77 (0.60 - 0.98)\*** |
|  | SC | 386<br>(15%) | 1,374<br>(15%) | 89 (16%) | 0.76<br>(0.60 - 0.95) | 0.81 (0.62 - 1.06) |
|  | ST | 502<br>(19%) | 1,606<br>(18%) | 134 (25%) | 0.74<br>(0.59 - 0.92) | 0.78 (0.60 - 1.02) |
|  | VJ/NT/Others | 335<br>(13%) | 1,082<br>(12%) | 54 (9.9%) | 0.66<br>(0.52 - 0.83) | **0.63 (0.48 - 0.83)\*** |
| **Marital status** | Currently married | 136<br>(5.3%) | 2,184<br>(25%) | 172 (32%) | 1 (ref) |  |
|  | Never married | 2,444<br>(95%) | 6,649<br>(75%) | 365 (67%) | 1.85<br>(0.92 - 3.57) |  |
|  | Divorced/ Separated | 0 (0%) | 41<br>(0.5%) | 6 (1.1%) | 0.28<br>(0.25 - 0.31) |  |
| **Occupation** | Farmer | 71<br>(2.8%) | 337<br>(3.8%) | 35 (6.4%) | 1 (ref) |  |
|  | Homemaker | 47<br>(1.8%) | 1,525<br>(17%) | 90 (17%) | 1.89<br>(1.45 - 2.46) |  |
|  | Labourer | 288<br>(11%) | 1,205<br>(14%) | 175 (32%) | 1.07<br>(0.82 -<br>1.39) |  |
|  | Service | 129<br>(5.0%) | 611<br>(6.9%) | 44 (8.1%) | 0.87<br>(0.65 -<br>1.16) |  |
|  | Student | 1,986<br>(77%) | 4,969<br>(56%) | 185 (34%) | 0.43<br>(0.34 -<br>0.54) |  |
|  | Unemployed | 59<br>(2.3%) | 227<br>(2.6%) | 14 (2.6%) | 0.69<br>(0.49 -<br>0.99) |  |
| **Education** | No or primary schooling | 1,398<br>(54%) | 2,708<br>(31%) | 135 (25%) | 1 (ref) | 1 (ref) |
|  | Secondary | 540<br>(21%) | 2,081<br>(23%) | 116 (21%) | 1.94<br>(1.74 -<br>2.16) | **1.64 (1.44**<br>**- 1.86)\*** |
|  | Higher Secondary | 460<br>(18%) | 2,914<br>(33%) | 186 (34%) | 2.95<br>(2.66 -<br>3.28) | **1.87 (1.65**<br>**- 2.13)\*** |
|  | Graduate and above | 179<br>(6.9%) | 1,168<br>(13%) | 106 (20%) | 3.41<br>(2.95 -<br>3.96) | **1.83 (1.51**<br>**- 2.22)\*** |
| **Wealth index** | Lowest | 275<br>(12%) | 804<br>(11%) | 72 (14%) | 1 (ref) | 1 (ref) |
|  | Second | 486<br>(22%) | 1,351<br>(19%) | 95 (19%) | 0.90<br>(0.76 -<br>1.05) | 0.92 (0.77<br>- 1.09) |
|  | Middle | 477<br>(21%) | 1,536<br>(22%) | 118 (23%) | 1.04<br>(0.89 -<br>1.22) | 0.94 (0.79<br>- 1.12) |
|  | Fourth | 478<br>(21%) | 1,729<br>(24%) | 113 (22%) | 1.11<br>(0.94 -<br>1.30) | 0.98 (0.82<br>- 1.16) |
|  | Highest | 512<br>(23%) | 1,709<br>(24%) | 107 (21%) | 1.03<br>(0.88 -<br>1.20) | 0.88 (0.74<br>- 1.05) |
| ***Smokeless***<br>***tobacco***<br>***consumers*** | No | 2266 | 7492 | 399 | 1 (ref) | 1 (ref) |
|  | Yes | 314<br>(12%) | 1,382<br>(16%) | 144 (27%) | 1.55<br>(1.38 -<br>1.75) | **1.50 (1.28**<br>**- 1.77)\*** |
| ***Alcohol***<br>***consumers***<br>***(past 30 days)*** | No | 2565 | 8689 | 527 | 1 (ref) | 1 (ref) |
|  | Yes | 15<br>(0.6%) | 185<br>(2.1%) | 16 (2.9%) | 2.52<br>(1.81 -<br>3.49) | **1.76 (1.17**<br>**- 2.67)\*** |
| ***Salt***<br>***consumption***<br>***(grams per***<br>***person per***<br>***day)*** | ≤ 5 grams | 121<br>(4.7%) | 329<br>(3.7%) | 36 (6.6%) | 1 (ref) |  |
|  | > 5 grams | 2,459<br>(95%) | 8,545<br>(96%) | 507 (93%) | 1.05<br>(0.86 -<br>1.30) |  |
| ***Sugar***<br>***consumption*** | ≤ 50 grams | 2,032<br>(79%) | 6,322<br>(71%) | 355 (65%) | 1 (ref) | 1 (ref) |
| *(grams per person per day)* | > 50 grams | 548<br>(21%) | 2,552<br>(29%) | 188 (35%) | 1.50<br>(1.37 - 1.65) | **1.47 (1.32 - 1.63)\*** |
| ***Fruit and vegetable servings per day*** | ≥ 5 servings per day | 3 (0.1%) | 3<br>(<0.1%) | 0 (0%) | 1 (ref) |  |
|  | < 5 servings per day | 2,571<br>(100%) | 8,866<br>(100%) | 543<br>(100%) | 3.75<br>(0.73 - 20.1) |  |
| ***Adequately physically active*** | No | 1985 | 5710 | 410 |  |  |
|  | Yes | 595<br>(23%) | 3,164<br>(36%) | 133 (24%) |  |  |
| ***BMI categories<sup>[18]</sup>*** | Normal | 1,586<br>(62%) | 4,902<br>(56%) | 289 (54%) | 1 (ref) | 1 (ref) |
|  | Severe Underweight | 205<br>(8.0%) | 658<br>(7.6%) | 34 (6.3%) | 1.01<br>(0.86 - 1.18) | 1.01 (0.84 - 1.20) |
|  | Underweight | 458<br>(18%) | 1,726<br>(20%) | 87 (16%) | 1.15<br>(1.03 -<br>1.28) | 1.04 (0.92<br>- 1.18) |
|  | Overweight | 179<br>(7.0%) | 710<br>(8.2%) | 63 (12%) | 1.39<br>(1.19 -<br>1.63) | 1.01 (0.85<br>- 1.21) |
|  | Obesity | 143<br>(5.6%) | 695<br>(8.0%) | 63 (12%) | 1.65<br>(1.40 -<br>1.95) | 0.99 (0.82<br>- 1.19) |
| **Hypertension**<br>[19] | Normal | 1,495<br>(58%) | 6,025<br>(68%) | 274 (50%) | 1 (ref) |  |
|  | Elevated | 270<br>(10%) | 722<br>(8.1%) | 33 (6.1%) | 0.70<br>(0.61 -<br>0.81) |  |
|  | HTN1 | 745<br>(29%) | 1,949<br>(22%) | 208 (38%) | 0.84<br>(0.76 -<br>0.93) |  |
|  | HTN2 | 69<br>(2.7%) | 177<br>(2.0%) | 28 (5.2%) | 0.98<br>(0.74 -<br>1.31) |  |
| ***Diabetes***<br>***Mellitus***<br>***status*** | Normal | 344<br>(93%) | 1,073<br>(94%) | 87 (97%) | 1 (ref) |  |
|  | Pre-diabetic | 19<br>(5.2%) | 58<br>(5.1%) | 2 (2.2%) | 0.85<br>(0.53 -<br>1.38) |  |
|  | Diabetic | 5 (1.4%) | 10<br>(0.9%) | 1 (1.1%) | 0.70<br>(0.25 -<br>2.10) |  |
| ***Depression*** | No | 2490 | 6011 | 198 | 1 (ref) | 1 (ref) |
|  | Yes | 90<br>(3.5%) | 2,863<br>(32%) | 345 (64%) | 8.41<br>(7.37 -<br>9.61) | **5.15 (4.39**<br>**- 6.07)\*** |
| ***Anxiety*** | No | 2546 | 7111 | 423 | 1 (ref) | 1 (ref) |
|  | Yes | 34<br>(1.3%) | 1,763<br>(20%) | 120 (22%) | 4.09<br>(3.60 -<br>4.66) | 0.97 (0.82<br>- 1.15) |
| No. Obs. |  |  | 9,711 |  |  |  |
| The model for stress shows odds adjusted for age category, sex, caste, education, wealth index, smokeless tobacco, alcohol in past 30 days, sugar consumption (grams per person per day), BMI categories, depression and anxiety |  |  |  |  |  |  |
| OR = Odds Ratio, aOR= adjusted Odds Ratio CI = Confidence Interval, OBC = Other backward class, SC = Scheduled caste, ST = Scheduled tribe, VJ/NT/Others = Vimukt jati/Nomadic Tribe/Others, BMI = Body Mass Index, HTN = Hypertension, TG = triglycerides , LDL = low density Lipoprotein, VLDL = Very low density Lipoprotein, HDL = high density Lipoprotein<br>*Indicates significant Odds ratios |  |  |  |  |  |  |

As mentioned in Table 3, compared to the 10-20 years age group, those in the 20-30 age group have 1.85 times significant odds of reporting high perceived stress than reporting low or moderate stress combined. Males had significantly lower odds (aOR = 0.37) of high perceived stress than combined odds of feeling low or moderate stress when compared to females. Those from OBC caste had lower odds (aOR = 0.77) of perceiving high stress than moderate and low stress when compared to open caste. VJ/NT/Others caste when compared to open caste also had a significant protective role against high perceived stress than moderate and low stress combined. (aOR = 0.63). Those with higher levels of education when compared to those with no or primary education had higher odds (aOR of those secondary education = 1.64, aOR of those with higher secondary = 1.87, aOR of those with graduate degree or higher = 1.83) of perceiving high stress than feeling moderate and low stress. Current smokeless tobacco consumers had 1.50 times significant odds of perceiving higher stress than combined odds of low and moderate stress when compared to those who didn’t consume smokeless tobacco. Respondents who reported alcohol consumption in the past 30 days had 1.76 times more significant odds of experiencing high stress than combined perception of moderate and low stress than those who didn’t consume alcohol. Participants who consumed sugar more than 50 grams per day had 1.47 times odds of perceiving higher stress than combined odds of low and moderate stress when compared to those who consumed sugar within recommended limit. Participants with reported depression had 5.15 times significant odds of reporting higher stress than combined odds of low and moderate stress when compared to those who weren’t depressed.

We have presented the predictors of smokeless tobacco consumption in table 4. The variables considered in the best fitting model (Nagelkerke’s R² = 0.5355) are mentioned in the footnote of table 4.

**Table 4:** Predictors for consumption of smokeless tobacco (Logistic regression model)

| Characteristic (N) |  | Smokeless tobacco non-consumers | Smokeless tobacco consumers | OR (95% CI) | aOR (95% CI) |
| --- | --- | --- | --- | --- | --- |
| **Age categories (11,997)** | 10-20 | 5,916 (58%) | 282 (15%) | 1 (ref) | 1 (ref) |
|  | 20-30 | 4,241 (42%) | 1,558 (85%) | 7.71 (6.76 - 8.82) | **3.09 (2.40 - 3.98)\*** |
| **Gender (11,997)** | Female | 5,741 (57%) | 169 (9.2%) | 1 (ref) | 1 (ref) |
|  | Male | 4,416 (43%) | 1,671 (91%) | 12.9 (11.0 - 15.2) | **15.4 (11.4 - 20.9)\*** |
| **Caste (11,997)** | Open | 441 (4.3%) | 59 (3.2%) | 1 (ref) | 1 (ref) |
|  | OBC | 5,202 (51%) | 733 (40%) | 1.05 (0.80 - 1.41) | 0.94 (0.63 - 1.42) |
|  | SC | 1,614 (16%) | 235 (13%) | 1.09 (0.81 - 1.49) | 1.07 (0.69 - 1.68) |
|  | ST | 1,699 (17%) | 543 (30%) | 2.39 (1.80 - 3.22) | **1.77 (1.16 - 2.75)\*** |
|  | VJ/NT/Others | 1,201 (12%) | 270 (15%) | 1.68 (1.25 - 2.29) | 1.42 (0.91 - 2.25) |
| **Marital Status**<br>**(11,997)** | Currently married | 2,014 (20%) | 478 (26%) | 1 (ref) | 1 (ref) |
|  | Divorced/<br>Separated | 37 (0.4%) | 10 (0.5%) | 1.14 (0.53 - 2.22) | 1.12 (0.35 - 3.06) |
|  | Never married | 8,106 (80%) | 1,352 (73%) | 0.70 (0.63 - 0.79) | **0.50 (0.39 - 0.63)\*** |
| **Occupation**<br>**(11,997)** | Farmer | 219 (2.2%) | 224 (12%) | 1 (ref) | 1 (ref) |
|  | Homemaker | 1,589 (16%) | 73 (4.0%) | 0.04 (0.03 - 0.06) | **0.24 (0.15 - 0.37)\*** |
|  | Labourer | 710 (7.0%) | 958 (52%) | 1.32 (1.07 - 1.63) | 1.09 (0.83 - 1.45) |
|  | Service | 524 (5.2%) | 260 (14%) | 0.49 (0.38 - 0.62) | **0.58 (0.42 - 0.78)\*** |
|  | Student | 6,871 (68%) | 269 (15%) | 0.04 (0.03 - 0.05) | **0.14 (0.10 - 0.20)\*** |
|  | Unemployed | 244 (2.4%) | 56 (3.0%) | 0.22 (0.16 - 0.31) | **0.49 (0.31 - 0.77)\*** |
| **Education**<br>**(11,991)** | No or Primary | 3,663 (36%) | 578 (31%) | 1 (ref) | 1 (ref) |
|  | Secondary | 2,155 (21%) | 582 (32%) | 1.71 (1.51 - 1.94) | 1.06 (0.86 - 1.30) |
|  | Higher Secondary | 2,983 (29%) | 577 (31%) | 1.23 (1.08 - 1.39) | 0.85 (0.69 - 1.05) |
|  | Graduate and above | 1,350 (13%) | 103 (5.6%) | 0.48 (0.39 - 0.60) | **0.28 (0.20 - 0.38)\*** |
| **Wealth index**<br>**(9,862)** | Lowest | 876 (10%) | 275 (19%) | 1 (ref) | 1 (ref) |
|  | Second | 1,604 (19%) | 328 (23%) | 0.65 (0.54 - 0.78) | **0.74 (0.57 - 0.95)\*** |
|  | Middle | 1,806 (21%) | 325 (23%) | 0.57 (0.48 - 0.69) | **0.56 (0.43 - 0.72)\*** |
|  | Fourth | 2,025 (24%) | 295 (20%) | 0.46 (0.39 - 0.56) | **0.61 (0.47 - 0.79)\*** |
|  | Highest | 2,110 (25%) | 218 (15%) | 0.33 (0.27 - 0.40) | **0.46 (0.34 - 0.61)\*** |
| **Alcohol consumption in the last 30 days (11,997)** | No | 10141 (99.8%) | 1640 (89%) | 1 (ref) | 1 (ref) |
|  | Yes | 16 (0.2%) | 200 (11%) | 77.3 (47.9 - 134) | **16.3 (8.43 - 35.0)\*** |
| **Salt consumption (Grams per person per day) (11,997)** | ≤ 5 grams | 420 (4.1%) | 66 (3.6%) | 1 (ref) | 1 (ref) |
|  | > 5 grams | 9,737 (95.9%) | 1,774 (96.4%) | 1.16 (0.90 - 1.52) | 1.16 (0.80 - 1.71) |
| **Sugar consumption (Grams per person per day) (11,997)** | ≤ 50 grams | 7,542 (74%) | 1,167 (63%) | 1 (ref) | 1 (ref) |
|  | > 50 grams | 2,615 (26%) | 673 (37%) | 1.66 (1.50 - 1.85) | **1.26 (1.07 - 1.49)\*** |
| **Fruit and vegetable** | ≥ 5 servings per day | 5 (<0.1%) | 1 (<0.1%) | 1 (ref) | 1 (ref) |
| ***servings per day (11,986)*** | < 5 servings per day | 10,143 (100%) | 1,837 (100%) | 0.91 (0.15 - 17.4) | 0.56 (0.05 - 16.3) |
| ***Adequate physical activity (11,997)*** | Yes | 2,896 (29%) | 996 (54%) | 1 (ref) | 1 (ref) |
|  | No | 7261 | 844 | 0.34 (0.31 - 0.37) | **0.82 (0.69 - 0.96)\*** |
| ***Stress (11,997)*** | Low stress | 2,266 (22%) | 314 (17%) | 1 (ref) | 1 (ref) |
|  | Moderate stress | 7,492 (74%) | 1,382 (75%) | 1.33 (1.17 - 1.52) | **1.33 (1.09 - 1.62)\*** |
|  | High perceived stress | 399 (3.9%) | 144 (7.8%) | 2.60 (2.08 - 3.25) | 0.97 (0.69 - 1.38) |
| ***BMI (11,798)<sup>[18]</sup>*** | Normal | 5,816 (58%) | 961 (52%) | 1 (ref) | 1 (ref) |
|  | Sev Underweight | 750 (7.5%) | 147 (8.0%) | 1.19 (0.98 - 1.43) | **1.41 (1.06 - 1.86)\*** |
|  | Underweight | 1,857 (19%) | 414 (23%) | 1.35 (1.19 - 1.53) | 1.17 (0.96 - 1.43) |
|  | Overweight | 806 (8.1%) | 146 (8.0%) | 1.10 (0.90 - 1.32) | **0.73 (0.55 - 0.96)\*** |
|  | Obesity | 736 (7.4%) | 165 (9.0%) | 1.36 (1.13 - 1.62) | 0.89 (0.68 - 1.15) |
| **Hypertension**<br>[19] **(11,995)** | Normal | 6,933 (68%) | 861 (47%) | 1 (ref) | 1 (ref) |
|  | Elevated | 758 (7.5%) | 267 (15%) | 2.84 (2.42 - 3.31) | 1.18 (0.93 - 1.51) |
|  | HTN1 | 2,268 (22%) | 634 (34%) | 2.25 (2.01 - 2.52) | 0.99 (0.82 - 1.18) |
|  | HTN2 | 196 (1.9%) | 78 (4.2%) | 3.20 (2.43 - 4.19) | 1.22 (0.80 - 1.83) |
| **Diabetes**<br>**(1,599)** | Normal | 1,274 (95%) | 230 (90%) | 1 (ref) |  |
|  | Pre-diabetic | 57 (4.2%) | 22 (8.6%) | 2.14 (1.26 - 3.52) |  |
|  | Diabetic | 13 (1.0%) | 3 (1.2%) | 1.28 (0.29 - 4.00) |  |
| **Depression**<br>**(11,997)** | No | 7361 | 1338 | 1 (ref) | 1 (ref) |
|  | Yes | 2,796 (28%) | 502 (27%) | 0.99 (0.88 -<br>1.10) | 1.12 (0.93 -<br>1.35) |
| **Anxiety**<br>**(11,997)** | No | 8449 | 1631 | 1 (ref) | 1 (ref) |
|  | Yes | 1,708 (17%) | 209 (11%) | 0.63 (0.54 -<br>0.74) | 1.12 (0.87 -<br>1.44) |
| No. Obs: 9,704 |  |  |  |  |  |
| The model for stress shows odds adjusted for age category, sex, caste, occupation, education, wealth index, alcohol in past 30 days, salt and sugar consumption (grams per person per day), fruit and vegetable consumption, adequate physical activity, stress, BMI categories, hypertension, depression and anxiety |  |  |  |  |  |
| OR = Odds Ratio, aOR= adjusted Odds Ratio CI = Confidence Interval, OBC = Other backward class, SC = Scheduled caste, ST = Scheduled tribe, VJ/NT/Others = Vimukt jati/Nomadic Tribe/Others, BMI = Body Mass Index, HTN = Hypertension<br>*Indicates significant Odds ratios |  |  |  |  |  |

Table 4 shows that compared to the 10-20 years age group, those in the 20-30 age group have 3.09 times significant odds of consuming smokeless tobacco. It was found that males have 15.4 times statistically significant odds of consuming smokeless tobacco than females. Respondents who identified as Scheduled Tribes have 1.77 times significant odds of consuming smokeless tobacco when compared to those from the Open category. Those respondents who were never married had significantly protective odds for consumption of smokeless tobacco (aOR = 050) than those who were married. Compared to respondents engaged in farming, those who were homemakers (aOR = 0.24), those in service (aOR = 0.58), students (aOR = 0.14) and those who were unemployed (aOR = 0.49) had significant odds of consuming smokeless tobacco and these associations were found to be statistically significant. Those who had education level of graduation and above (aOR = 0.28) as well as respondents belonging to the second (aOR = 0.74), middle (aOR = 0.56), fourth (aOR = 0.61) and highest strata (aOR = 0.46) based on wealth index had significantly lower odds of consuming smokeless tobacco. Respondents who consumed alcohol in the past 30 days have significantly higher odds (aOR = 16.3) of consuming smokeless tobacco than those who didn’t consume alcohol. For every gram per person per day increase in sugar intake, the odds of consuming smokeless tobacco significantly increase by 1.26 times. Those not indulging in adequate physical activity had lower odds of consuming smokeless tobacco (aOR = 0.82). Those experiencing moderate stress had 1.33 times significant odds of consuming smokeless tobacco than those reporting low stress. Respondents who were severely underweight had higher odds (aOR = 1.41) whereas those who were obese (aOR = 0.73) had lower odds for consumption of smokeless tobacco.

To study the predictors of physical activity a logistic regression model was constructed and the results are depicted in table 5. The final best fit model has a Nagelkerke R^2^ value of 0.2079.

**Table 5:**
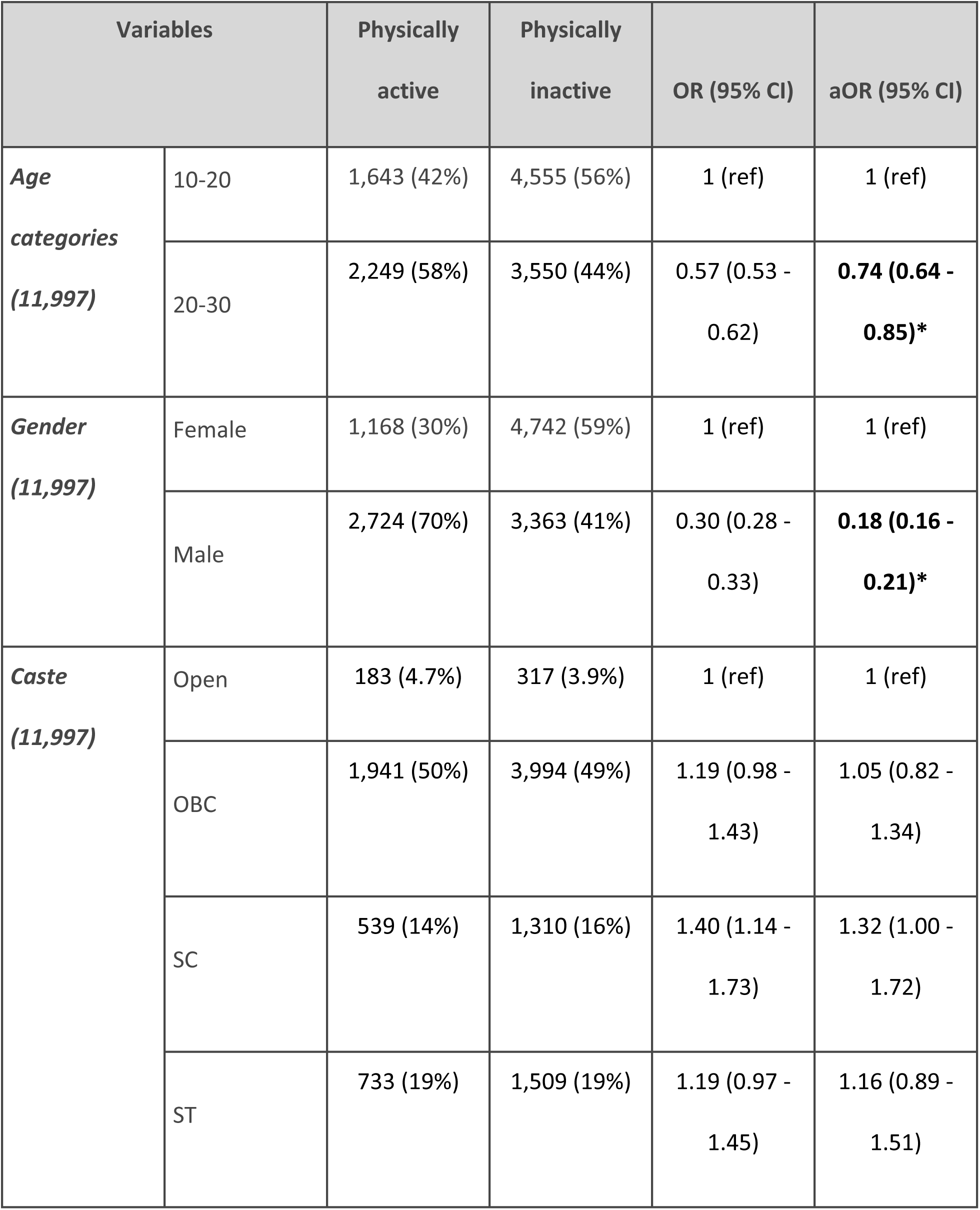
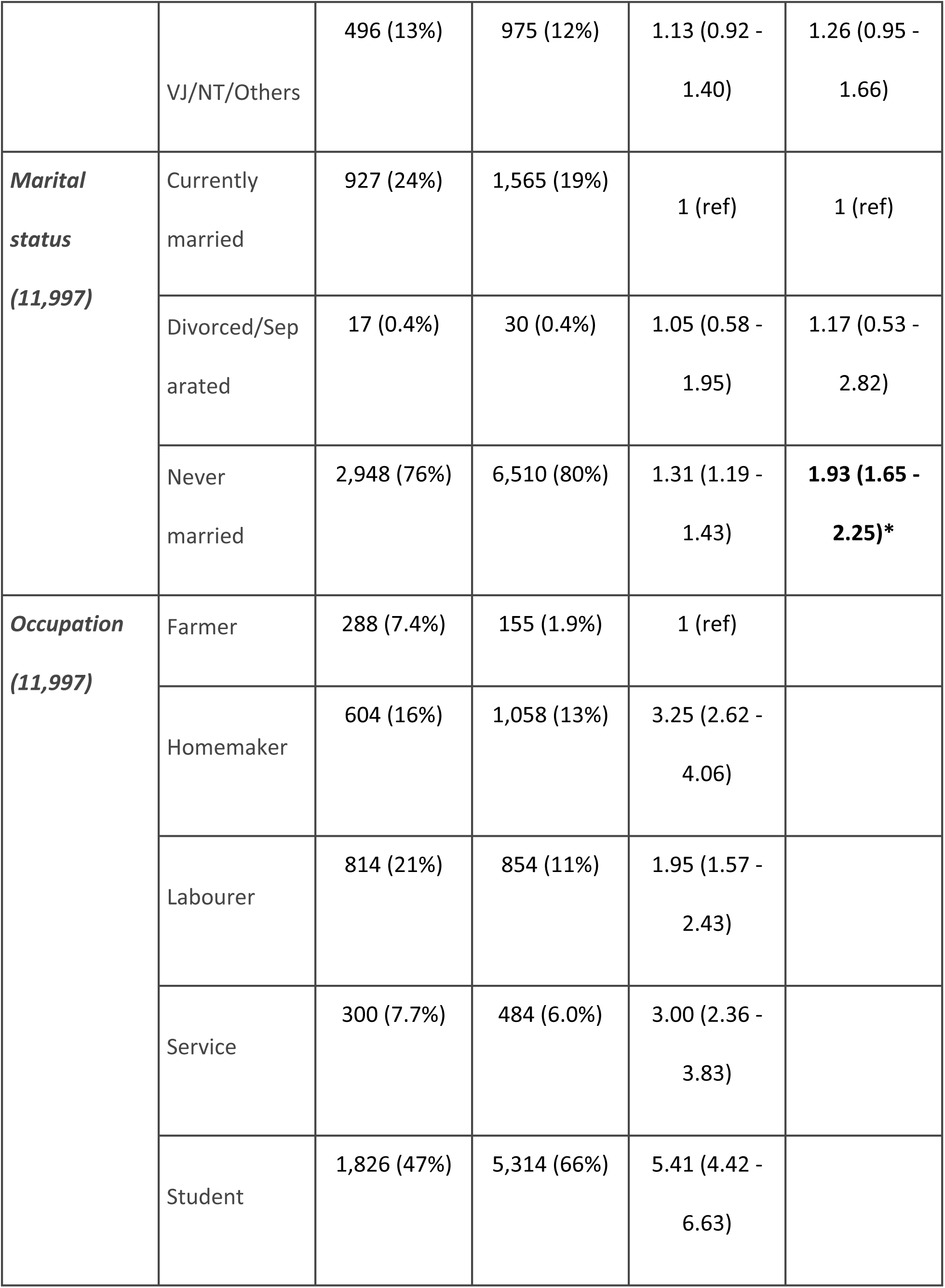
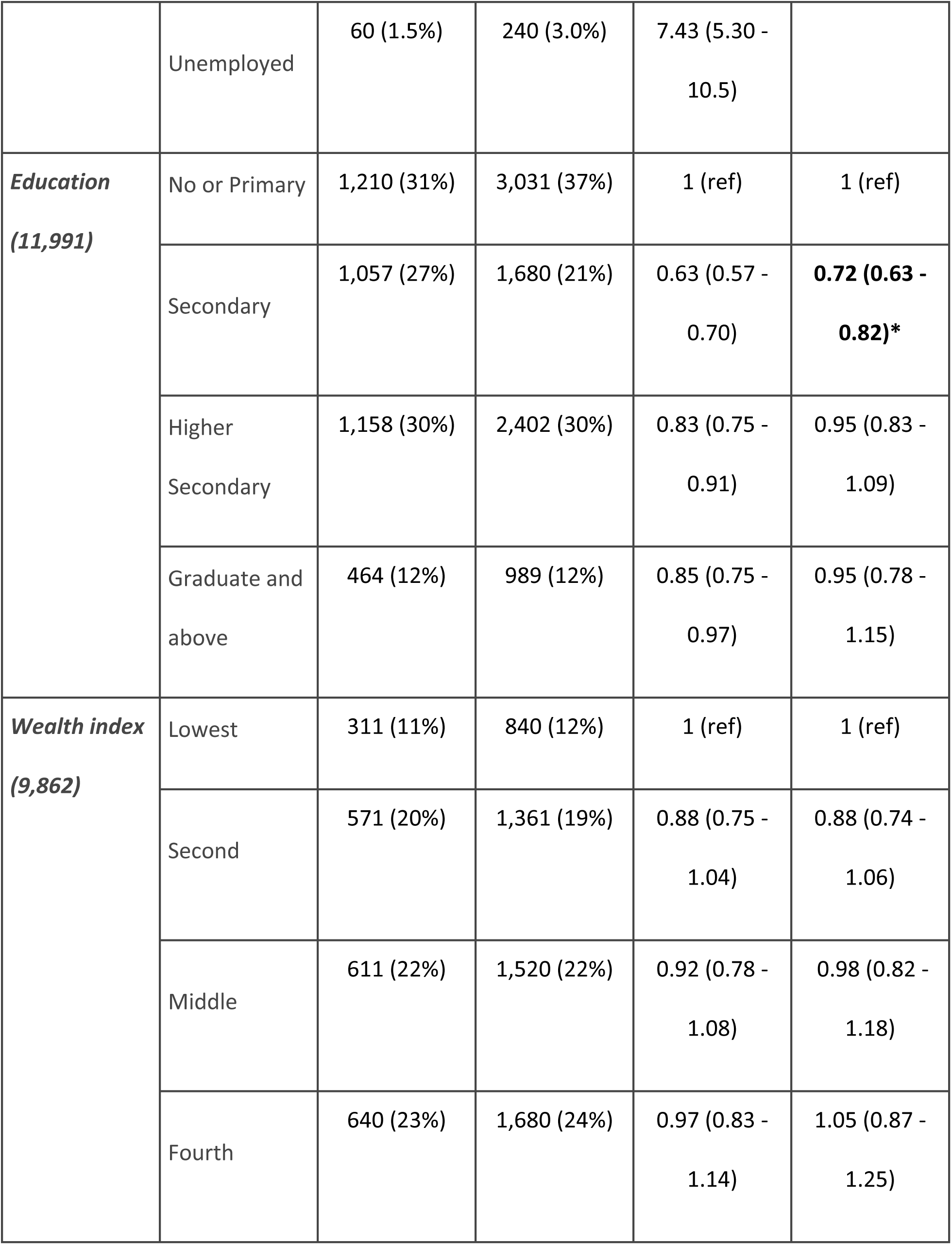
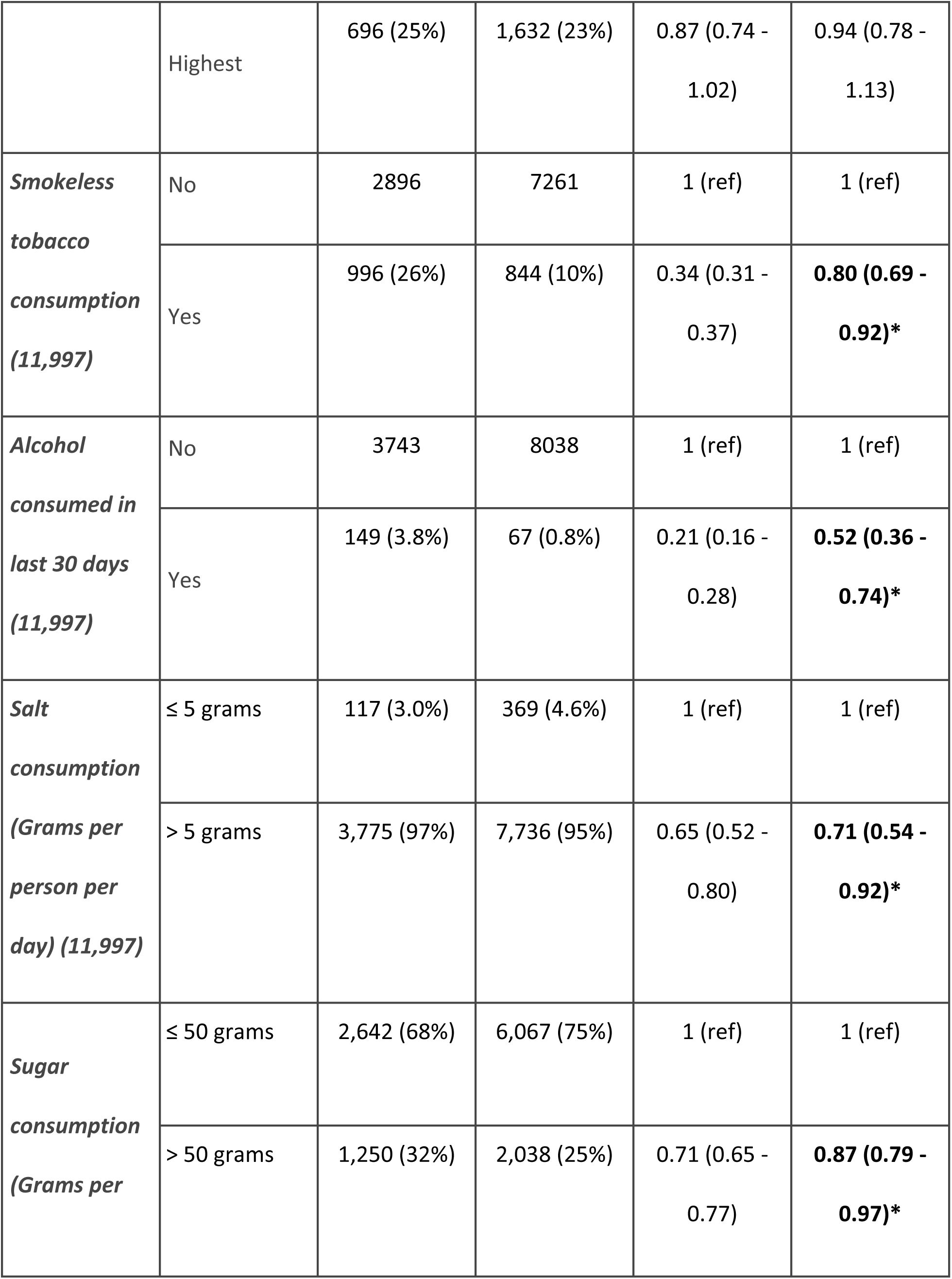
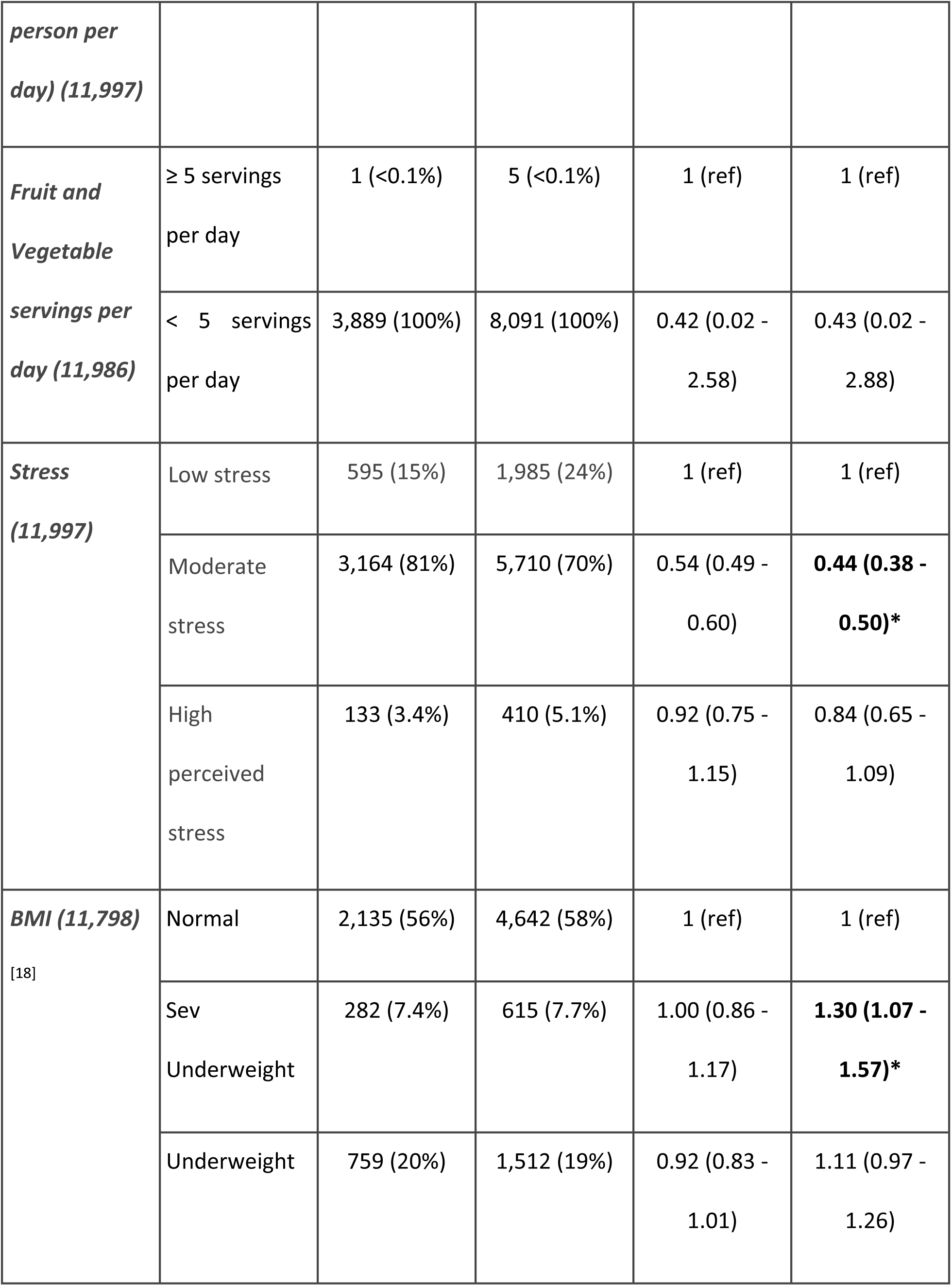
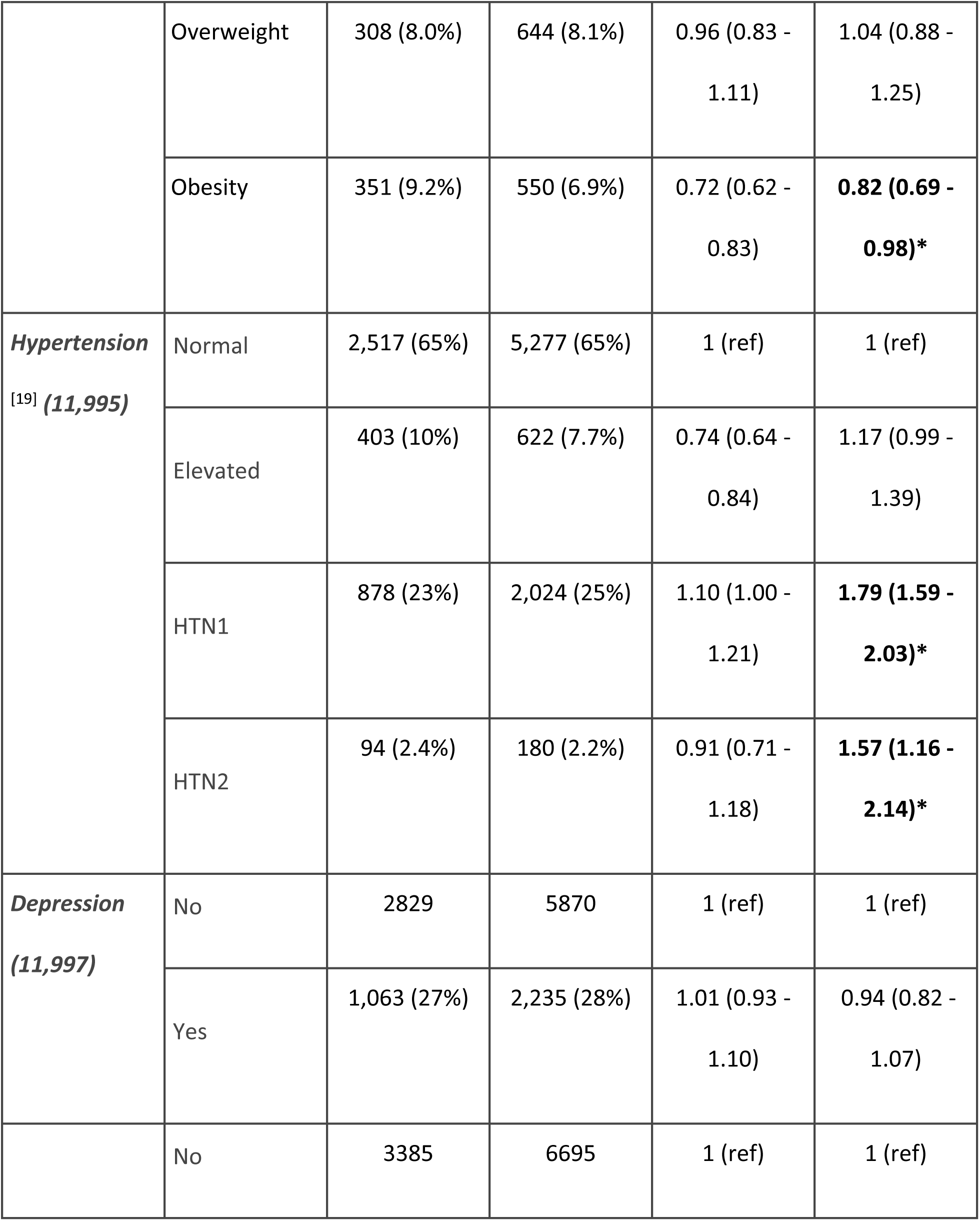
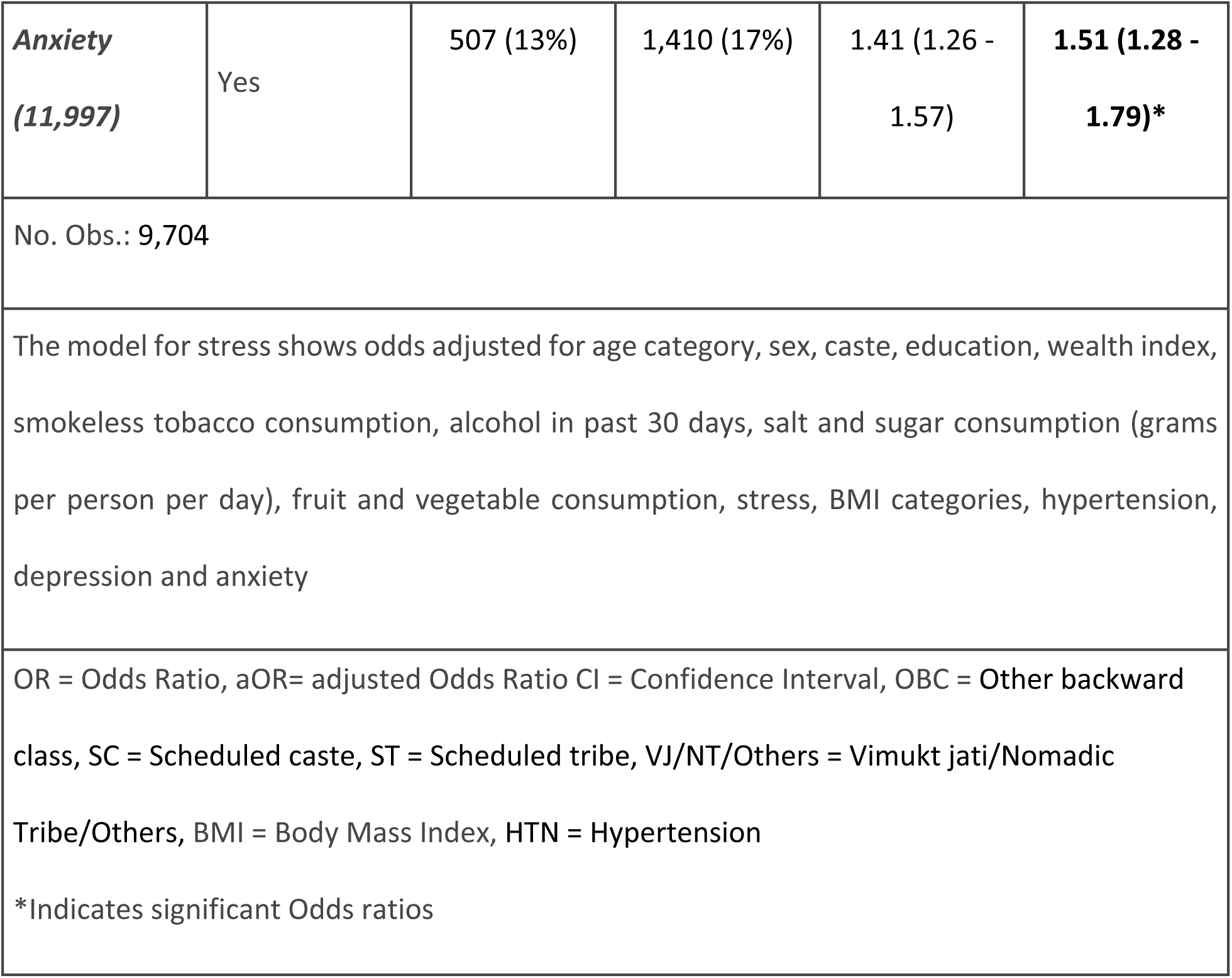
Predictors for inadequate physical activity (logistic regression model)

As seen in table 5, respondents in the age group 20-30 years had significantly lower odds of indulging in adequate physical activity (aOR = 0.74). It was found that males have 0.18 times significant odds of not indulging in physical activity compared to females. Respondents who were never married had 1.93 times more significant odds of not indulging in adequate physical activity than those who were married at the time of data collection. Those who had completed their secondary schooling had 0.72 times significant odds of not being adequately physically active compared to those who had no or primary schooling. Those who reported consuming smokeless tobacco had 0.80 times significant odds and those who consumed alcohol in last 30 days had 0.52 times significant odds of not being physically active than those who didn’t consume it. Those consuming excess salt (> 5 grams) and excess sugar (>50 grams) had 071 and 0.87 times significant odds of not indulging in adequate physical activity. Participants who reported moderate stress had 0.44 times significant odds of indulging in inadequate physical activity. Respondents who were severely underweight and obese had 1.3 and 0.82 times significant odds of being physically inactive. Participants classified as HTN grade I and II were found to have significantly higher odds of being physically inactive (aOR= 1.79 and 1.57 respectively). Those respondents who reported feeling anxious have 1.51 times significant odds to be physically inactive. (Table 5)

To understand the predictors of excess salt (Nagelkerke’s R² = 0.14) and excess sugar consumption (Nagelkerke’s R² = 0.1224), we developed a logistic regression model. Those from the second wealth quintile have 3.17 times the odds to have excess salt consumption compared to those who belong to the lowest wealth quintile. Respondents reporting excess sugar consumption have significantly higher odds (aOP = 4.94) of consuming an excess amount of salt. Adolescents who reported moderate levels of stress had significant odds of 1.97 times of consuming excess salt than those who had low stress. Respondents who had elevated blood pressure and those categorised as having pre-diabetes had significantly lower odds of consuming excess amounts of salt (aOR = 0.39 and 0.30 respectively) of consuming an excess salt. This model created for understanding predictors for excess salt consumption included gender, caste, marital status, wealth index, smokeless tobacco, alcohol and sugar consumption, fruit and vegetable servings, physical activity, stress, hypertension, depression, anxiety, TG, LDL, HDL, age, BMI categories, diabetes status. (table 6)

**Table 6:**
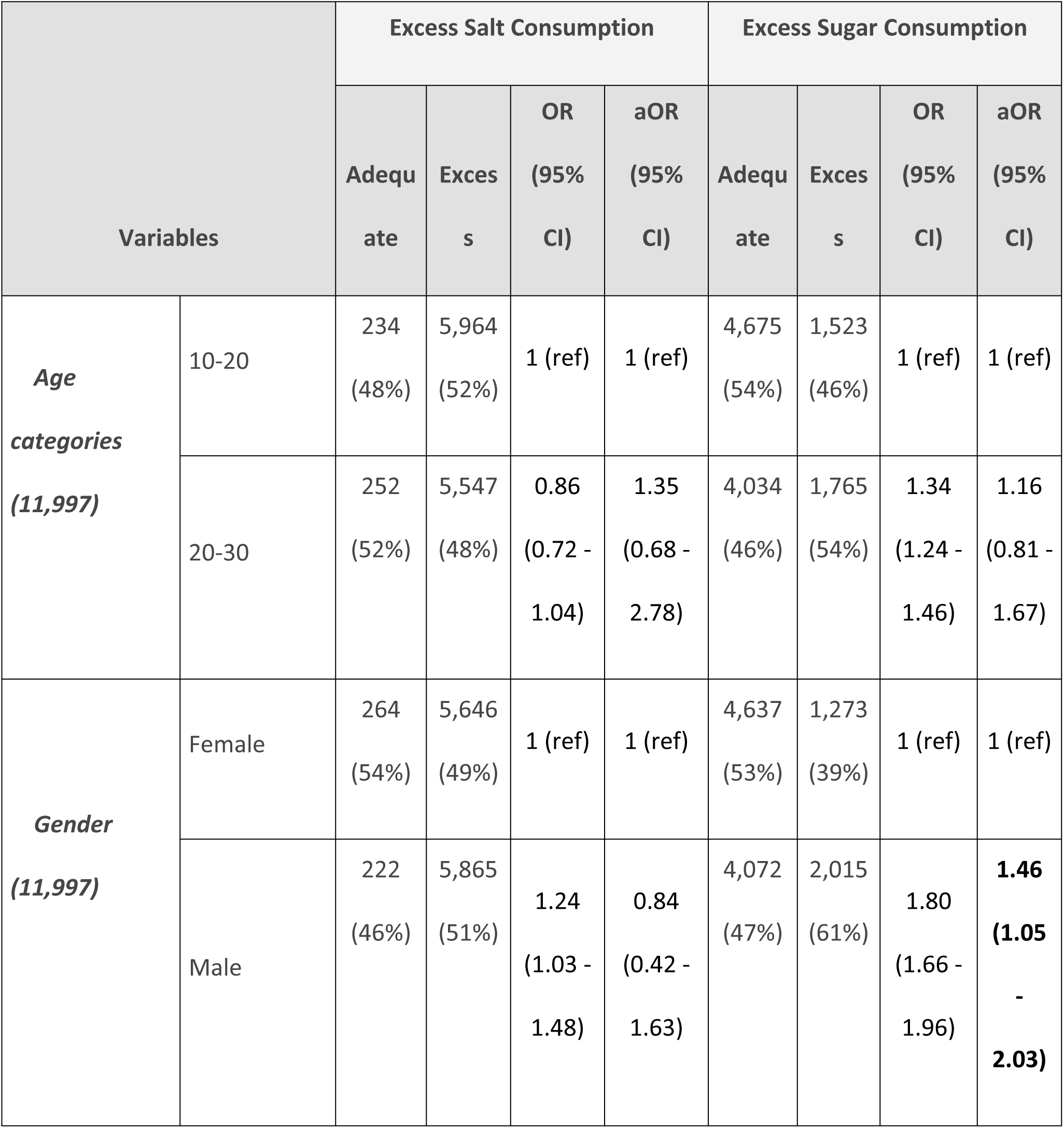
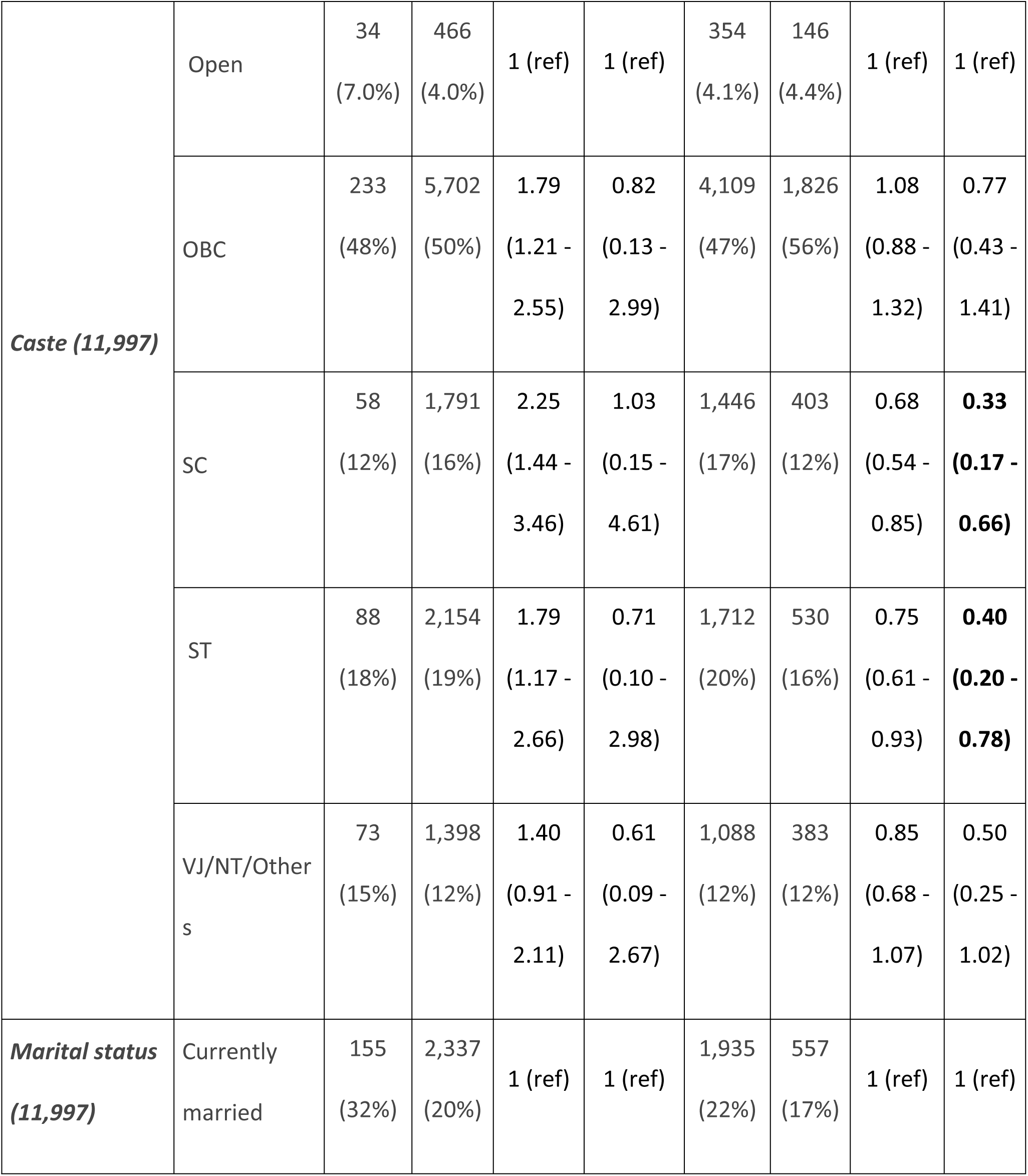
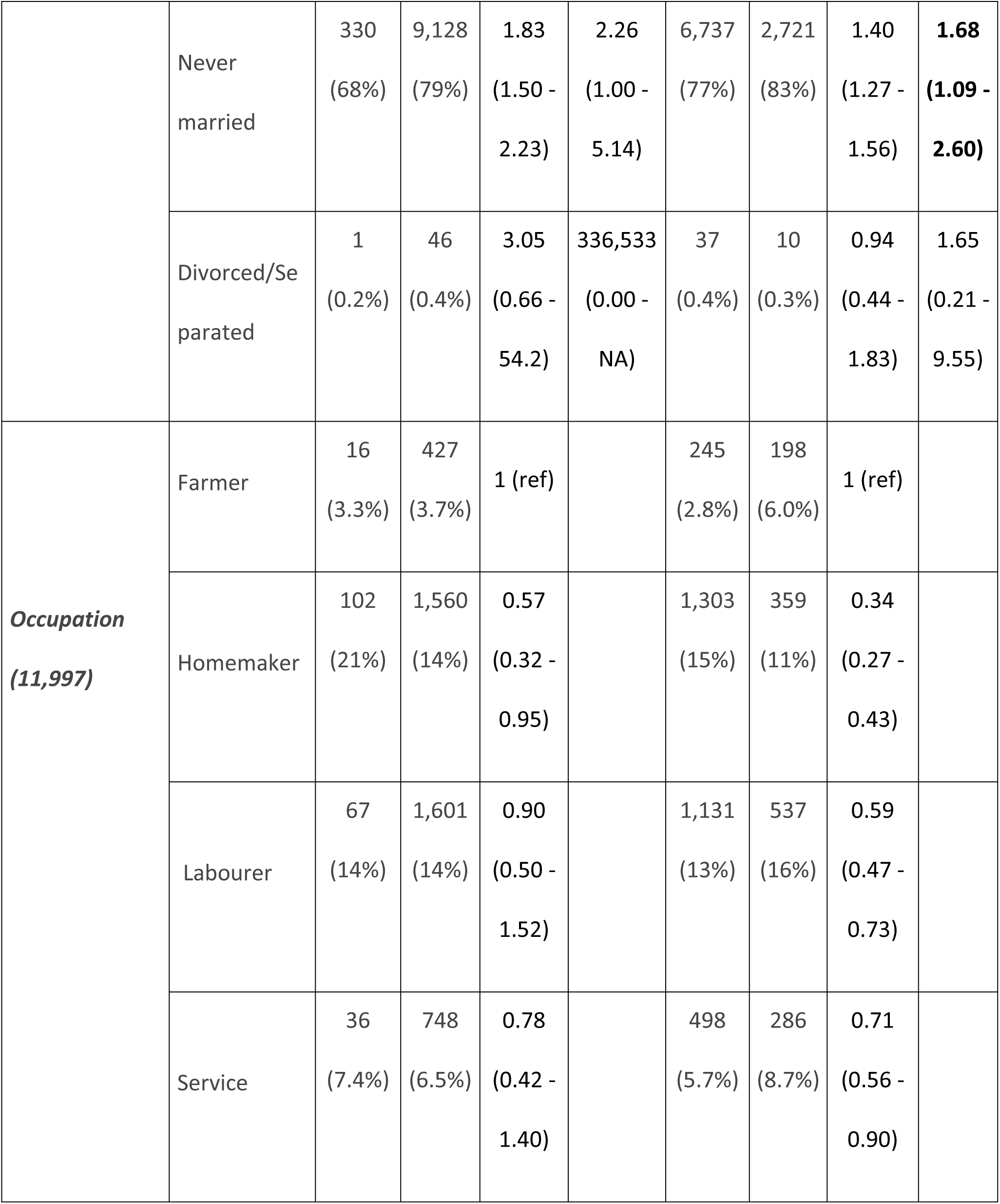
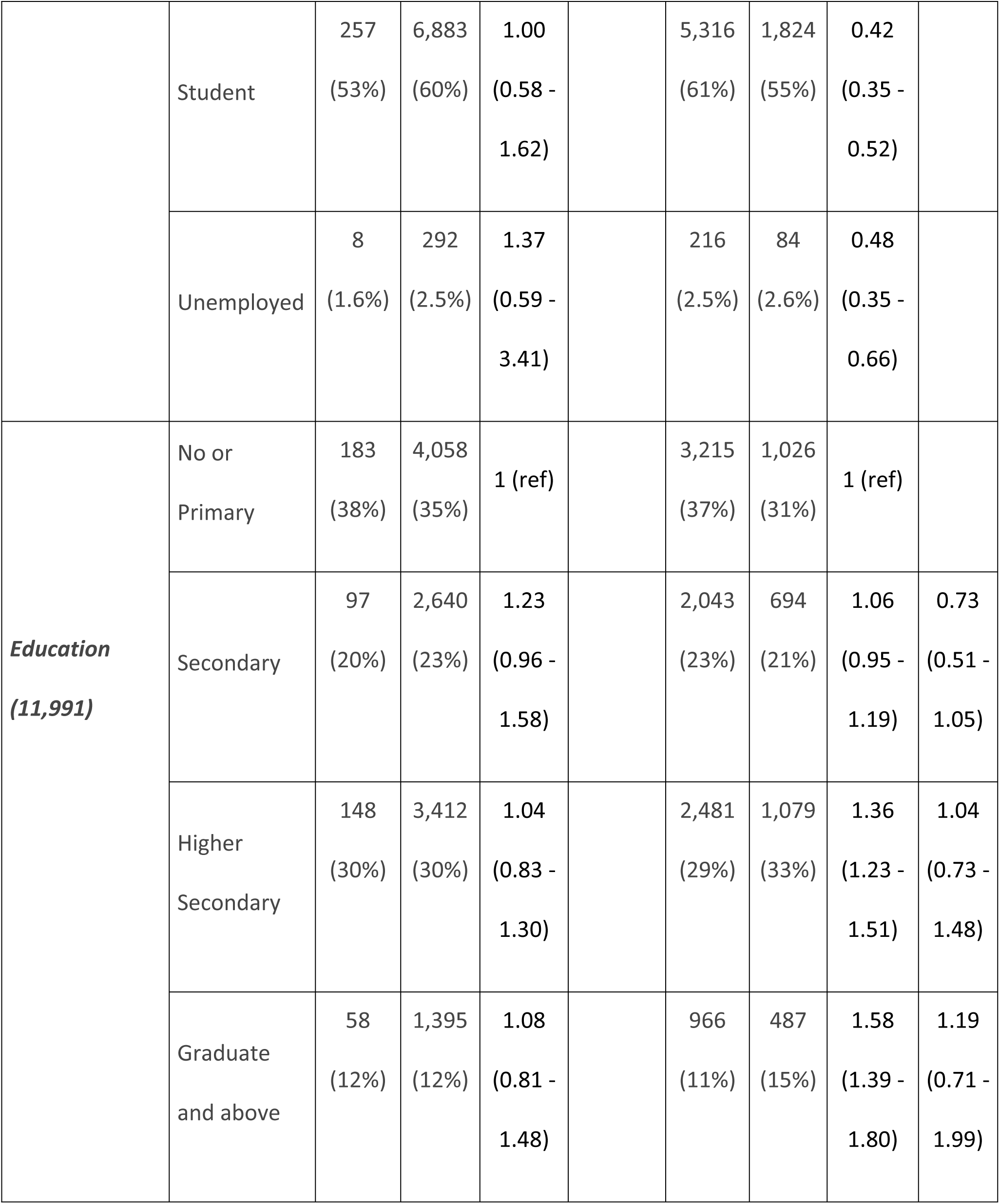
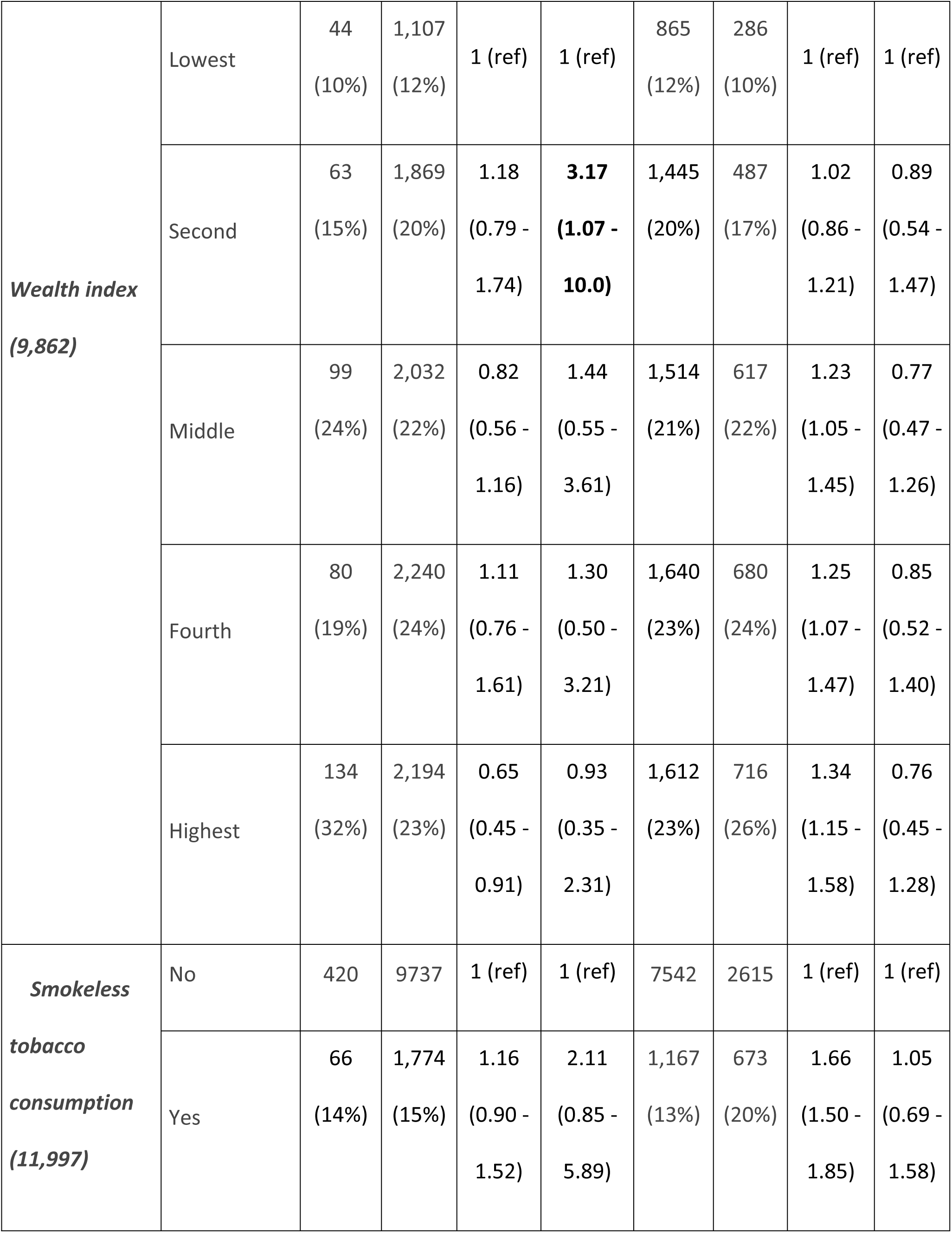
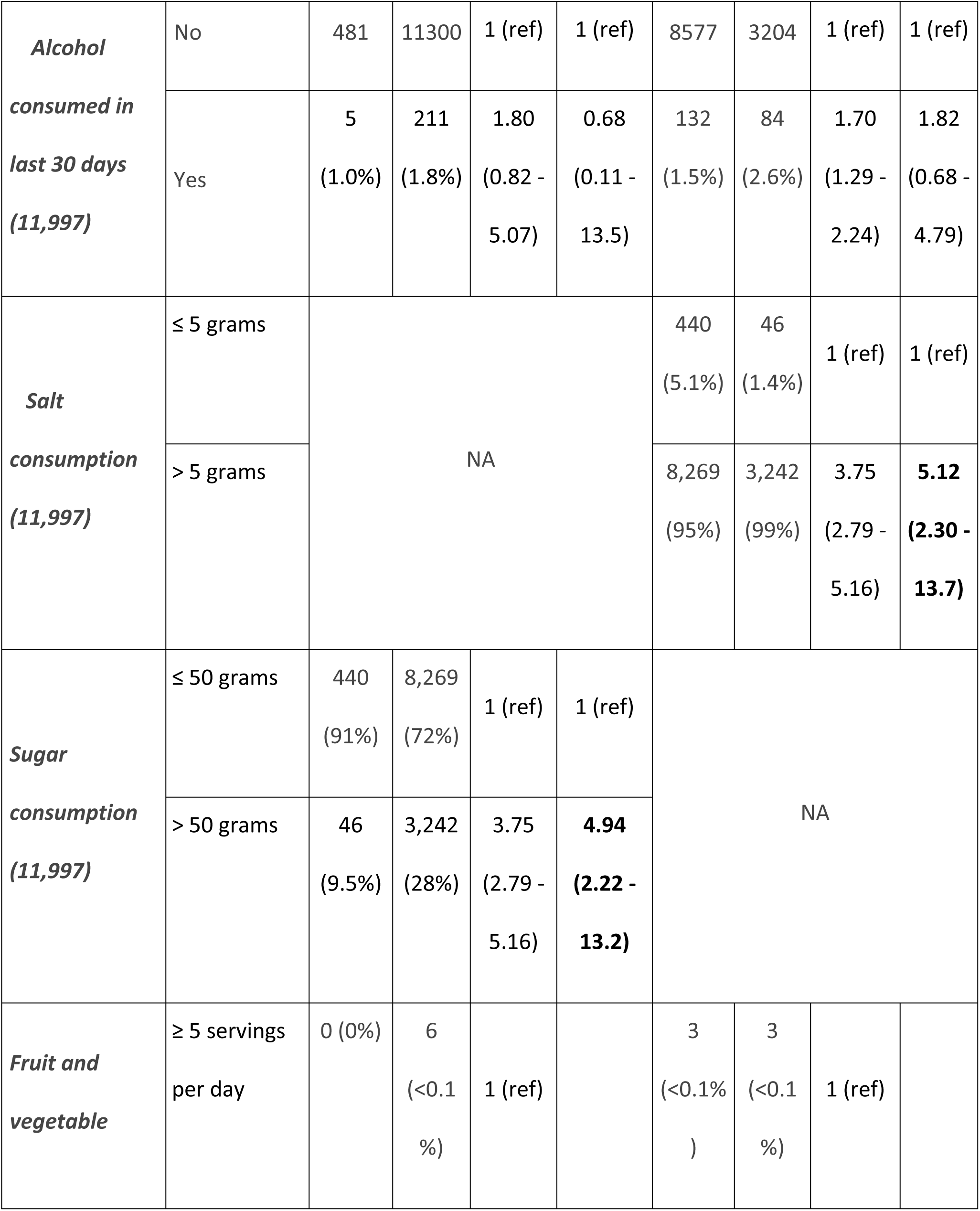
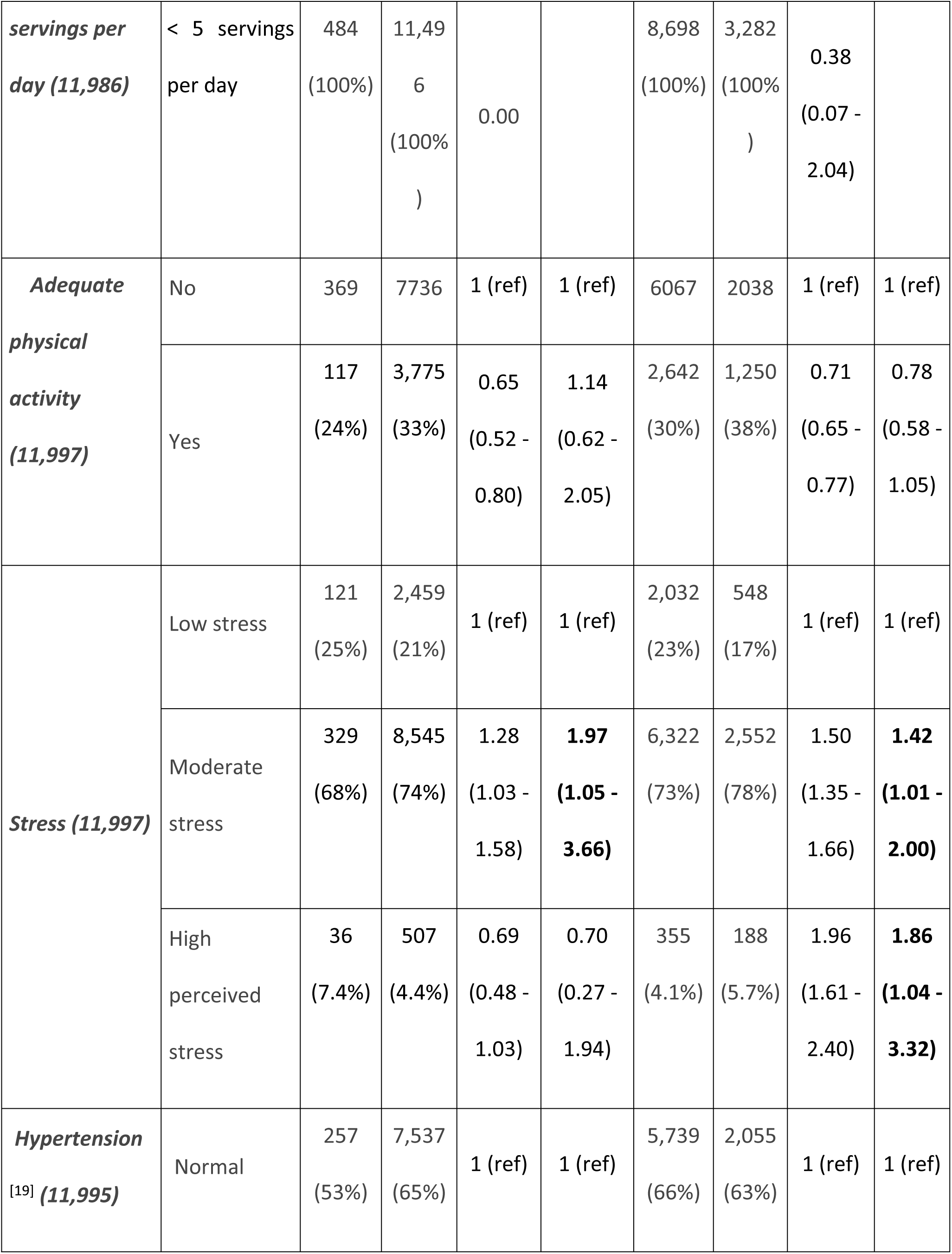
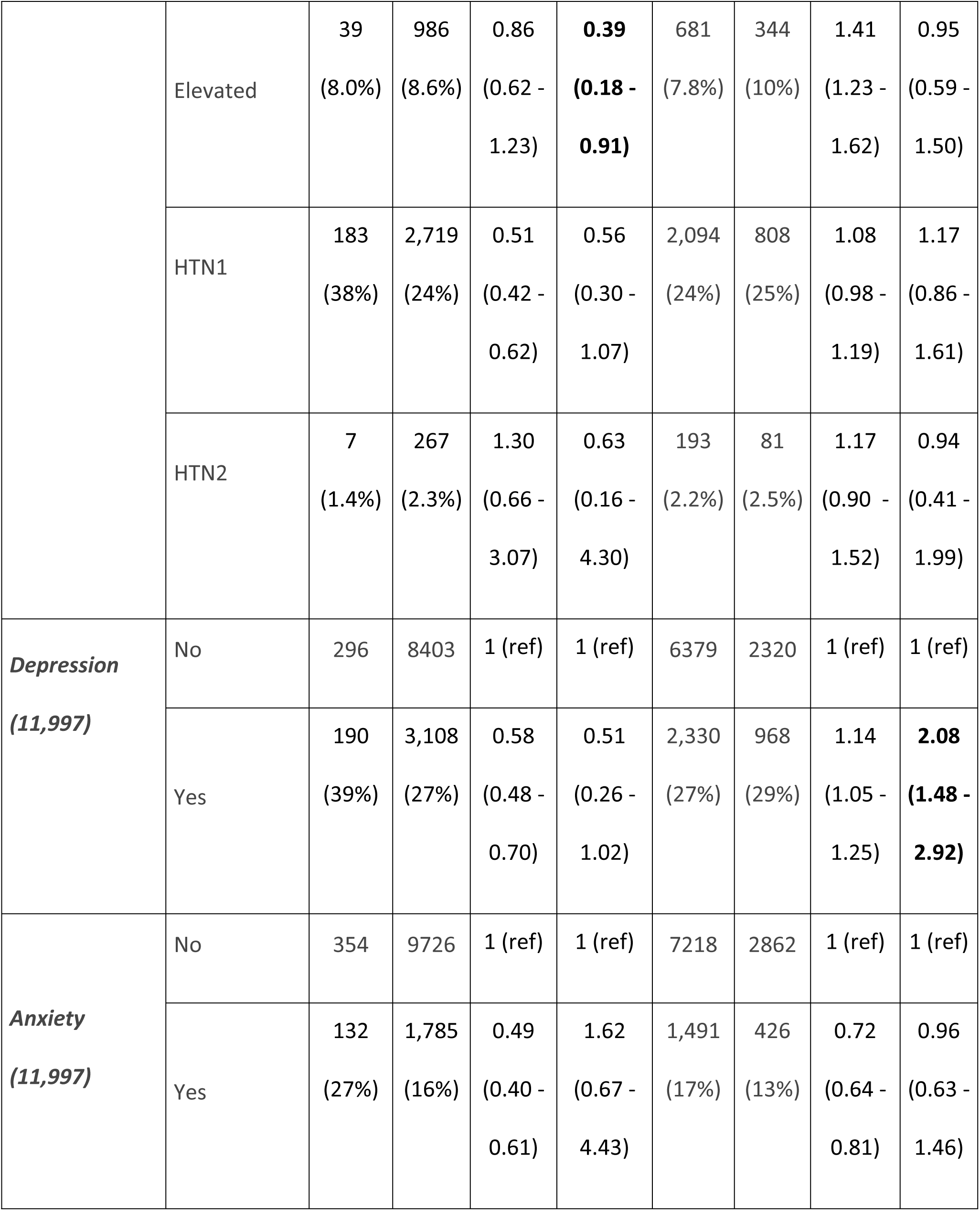
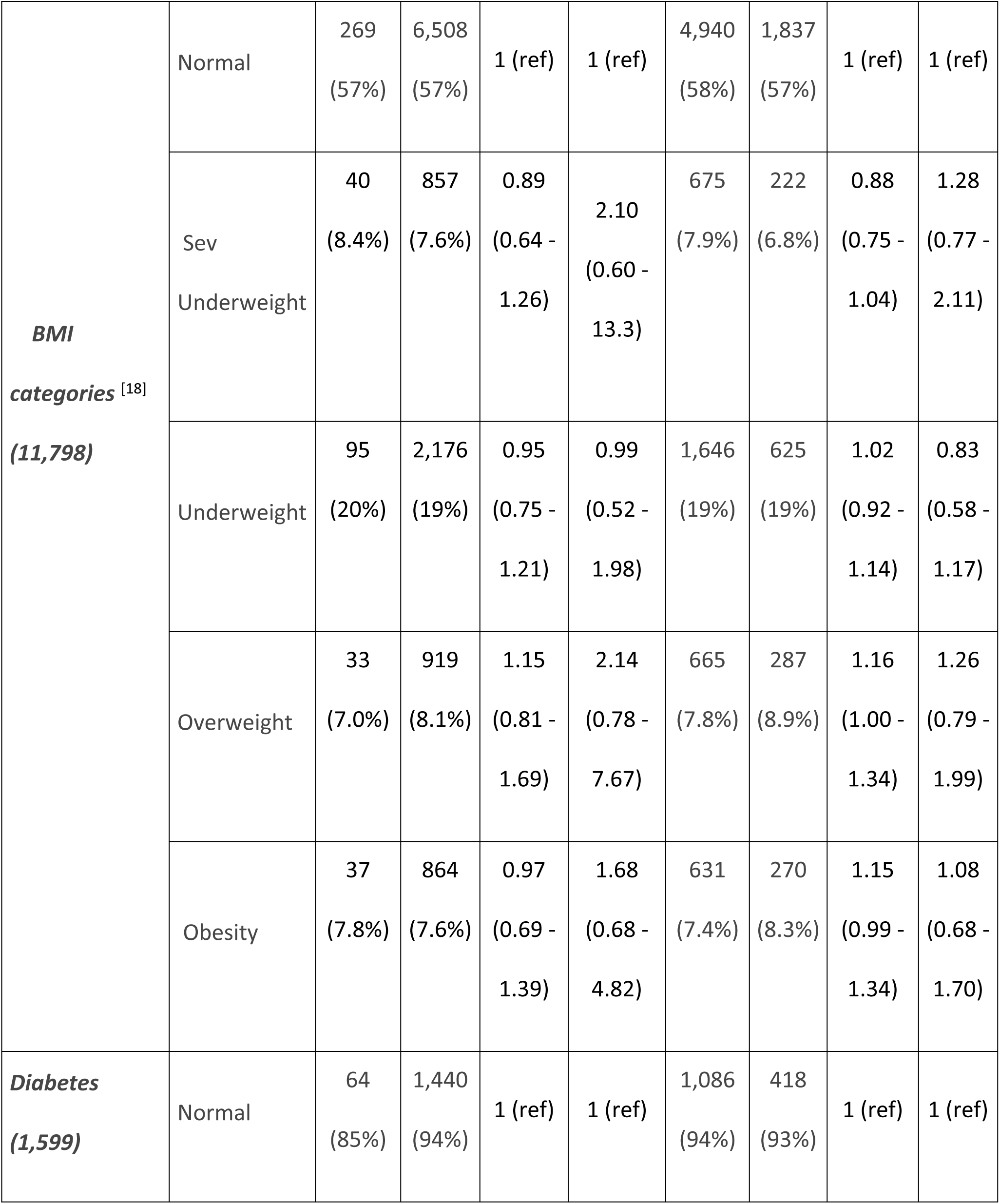
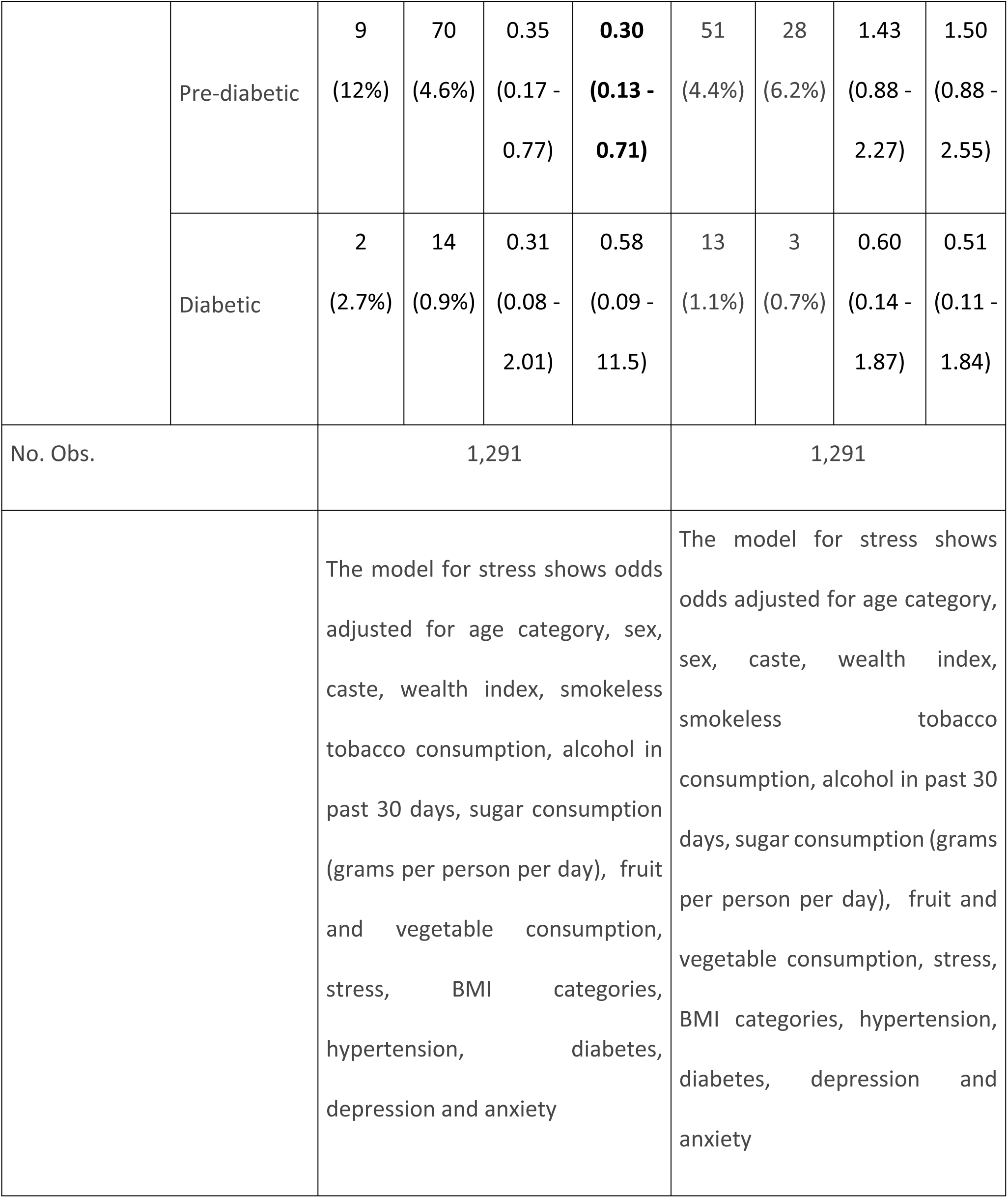
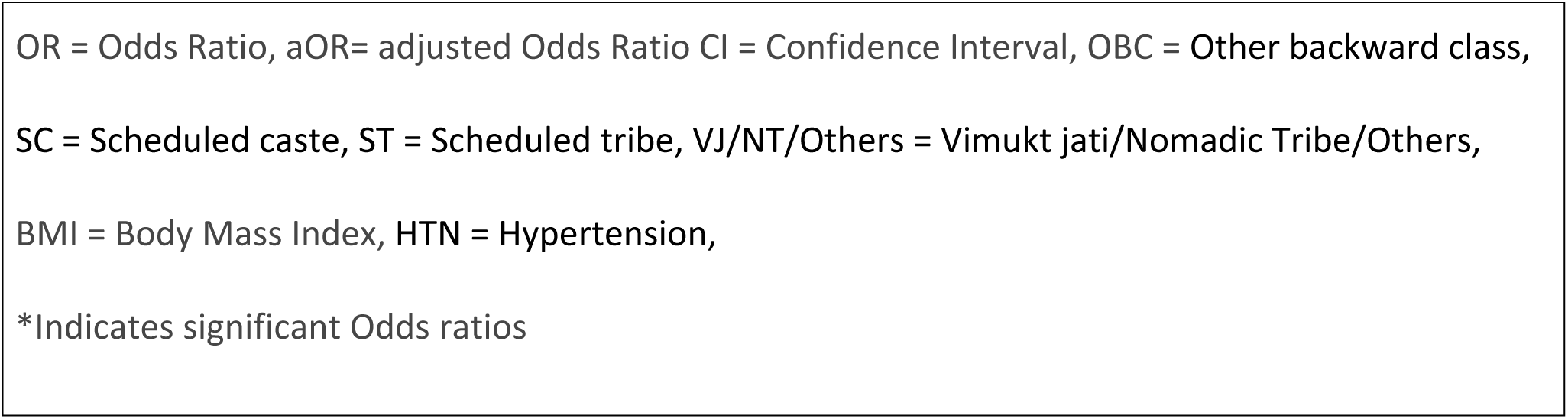
Predictors of excess salt and sugar consumption (Logistic regression)

| Variables |  | Excess Salt Consumption |  |  |  | Excess Sugar Consumption |  |  |  |
| --- | --- | --- | --- | --- | --- | --- | --- | --- | --- |
|  |  | Adequ<br>ate | Exces<br>s | OR<br>(95%<br>CI) | aOR<br>(95%<br>CI) | Adequ<br>ate | Exces<br>s | OR<br>(95%<br>CI) | aOR<br>(95%<br>CI) |
| **Age**<br>**categories**<br>**(11,997)** | 10-20 | 234<br>(48%) | 5,964<br>(52%) | 1 (ref) | 1 (ref) | 4,675<br>(54%) | 1,523<br>(46%) | 1 (ref) | 1 (ref) |
|  | 20-30 | 252<br>(52%) | 5,547<br>(48%) | 0.86<br>(0.72 -<br>1.04) | 1.35<br>(0.68 -<br>2.78) | 4,034<br>(46%) | 1,765<br>(54%) | 1.34<br>(1.24 -<br>1.46) | 1.16<br>(0.81 -<br>1.67) |
| **Gender**<br>**(11,997)** | Female | 264<br>(54%) | 5,646<br>(49%) | 1 (ref) | 1 (ref) | 4,637<br>(53%) | 1,273<br>(39%) | 1 (ref) | 1 (ref) |
|  | Male | 222<br>(46%) | 5,865<br>(51%) | 1.24<br>(1.03 -<br>1.48) | 0.84<br>(0.42 -<br>1.63) | 4,072<br>(47%) | 2,015<br>(61%) | 1.80<br>(1.66 -<br>1.96) | **1.46**<br>**(1.05**<br>**-**<br>**2.03)** |
| **Caste (11,997)** | Open | 34<br>(7.0%) | 466<br>(4.0%) | 1 (ref) | 1 (ref) | 354<br>(4.1%) | 146<br>(4.4%) | 1 (ref) | 1 (ref) |
|  | OBC | 233<br>(48%) | 5,702<br>(50%) | 1.79<br>(1.21 -<br>2.55) | 0.82<br>(0.13 -<br>2.99) | 4,109<br>(47%) | 1,826<br>(56%) | 1.08<br>(0.88 -<br>1.32) | 0.77<br>(0.43 -<br>1.41) |
|  | SC | 58<br>(12%) | 1,791<br>(16%) | 2.25<br>(1.44 -<br>3.46) | 1.03<br>(0.15 -<br>4.61) | 1,446<br>(17%) | 403<br>(12%) | 0.68<br>(0.54 -<br>0.85) | **0.33**<br>**(0.17 -**<br>**0.66)** |
|  | ST | 88<br>(18%) | 2,154<br>(19%) | 1.79<br>(1.17 -<br>2.66) | 0.71<br>(0.10 -<br>2.98) | 1,712<br>(20%) | 530<br>(16%) | 0.75<br>(0.61 -<br>0.93) | **0.40**<br>**(0.20 -**<br>**0.78)** |
|  | VJ/NT/Other<br>s | 73<br>(15%) | 1,398<br>(12%) | 1.40<br>(0.91 -<br>2.11) | 0.61<br>(0.09 -<br>2.67) | 1,088<br>(12%) | 383<br>(12%) | 0.85<br>(0.68 -<br>1.07) | 0.50<br>(0.25 -<br>1.02) |
| **Marital status**<br>**(11,997)** | Currently<br>married | 155<br>(32%) | 2,337<br>(20%) | 1 (ref) | 1 (ref) | 1,935<br>(22%) | 557<br>(17%) | 1 (ref) | 1 (ref) |
|  | Never married | 330<br>(68%) | 9,128<br>(79%) | 1.83<br>(1.50 - 2.23) | 2.26<br>(1.00 - 5.14) | 6,737<br>(77%) | 2,721<br>(83%) | 1.40<br>(1.27 - 1.56) | **1.68**<br>**(1.09 - 2.60)** |
|  | Divorced/Separated | 1<br>(0.2%) | 46<br>(0.4%) | 3.05<br>(0.66 - 54.2) | 336,533<br>(0.00 - NA) | 37<br>(0.4%) | 10<br>(0.3%) | 0.94<br>(0.44 - 1.83) | 1.65<br>(0.21 - 9.55) |
| **Occupation**<br>**(11,997)** | Farmer | 16<br>(3.3%) | 427<br>(3.7%) | 1 (ref) |  | 245<br>(2.8%) | 198<br>(6.0%) | 1 (ref) |  |
|  | Homemaker | 102<br>(21%) | 1,560<br>(14%) | 0.57<br>(0.32 - 0.95) |  | 1,303<br>(15%) | 359<br>(11%) | 0.34<br>(0.27 - 0.43) |  |
|  | Labourer | 67<br>(14%) | 1,601<br>(14%) | 0.90<br>(0.50 - 1.52) |  | 1,131<br>(13%) | 537<br>(16%) | 0.59<br>(0.47 - 0.73) |  |
|  | Service | 36<br>(7.4%) | 748<br>(6.5%) | 0.78<br>(0.42 - 1.40) |  | 498<br>(5.7%) | 286<br>(8.7%) | 0.71<br>(0.56 - 0.90) |  |
|  | Student | 257<br>(53%) | 6,883<br>(60%) | 1.00<br>(0.58 -<br>1.62) |  | 5,316<br>(61%) | 1,824<br>(55%) | 0.42<br>(0.35 -<br>0.52) |  |
|  | Unemployed | 8<br>(1.6%) | 292<br>(2.5%) | 1.37<br>(0.59 -<br>3.41) |  | 216<br>(2.5%) | 84<br>(2.6%) | 0.48<br>(0.35 -<br>0.66) |  |
| **Education**<br>**(11,991)** | No or<br>Primary | 183<br>(38%) | 4,058<br>(35%) | 1 (ref) |  | 3,215<br>(37%) | 1,026<br>(31%) | 1 (ref) |  |
|  | Secondary | 97<br>(20%) | 2,640<br>(23%) | 1.23<br>(0.96 -<br>1.58) |  | 2,043<br>(23%) | 694<br>(21%) | 1.06<br>(0.95 -<br>1.19) | 0.73<br>(0.51 -<br>1.05) |
|  | Higher<br>Secondary | 148<br>(30%) | 3,412<br>(30%) | 1.04<br>(0.83 -<br>1.30) |  | 2,481<br>(29%) | 1,079<br>(33%) | 1.36<br>(1.23 -<br>1.51) | 1.04<br>(0.73 -<br>1.48) |
|  | Graduate<br>and above | 58<br>(12%) | 1,395<br>(12%) | 1.08<br>(0.81 -<br>1.48) |  | 966<br>(11%) | 487<br>(15%) | 1.58<br>(1.39 -<br>1.80) | 1.19<br>(0.71 -<br>1.99) |
| **Wealth index**<br>**(9,862)** | Lowest | 44<br>(10%) | 1,107<br>(12%) | 1 (ref) | 1 (ref) | 865<br>(12%) | 286<br>(10%) | 1 (ref) | 1 (ref) |
|  | Second | 63<br>(15%) | 1,869<br>(20%) | 1.18<br>(0.79 -<br>1.74) | **3.17**<br>**(1.07 -**<br>**10.0)** | 1,445<br>(20%) | 487<br>(17%) | 1.02<br>(0.86 -<br>1.21) | 0.89<br>(0.54 -<br>1.47) |
|  | Middle | 99<br>(24%) | 2,032<br>(22%) | 0.82<br>(0.56 -<br>1.16) | 1.44<br>(0.55 -<br>3.61) | 1,514<br>(21%) | 617<br>(22%) | 1.23<br>(1.05 -<br>1.45) | 0.77<br>(0.47 -<br>1.26) |
|  | Fourth | 80<br>(19%) | 2,240<br>(24%) | 1.11<br>(0.76 -<br>1.61) | 1.30<br>(0.50 -<br>3.21) | 1,640<br>(23%) | 680<br>(24%) | 1.25<br>(1.07 -<br>1.47) | 0.85<br>(0.52 -<br>1.40) |
|  | Highest | 134<br>(32%) | 2,194<br>(23%) | 0.65<br>(0.45 -<br>0.91) | 0.93<br>(0.35 -<br>2.31) | 1,612<br>(23%) | 716<br>(26%) | 1.34<br>(1.15 -<br>1.58) | 0.76<br>(0.45 -<br>1.28) |
| **Smokeless**<br>**tobacco**<br>**consumption**<br>**(11,997)** | No | 420 | 9737 | 1 (ref) | 1 (ref) | 7542 | 2615 | 1 (ref) | 1 (ref) |
|  | Yes | 66<br>(14%) | 1,774<br>(15%) | 1.16<br>(0.90 -<br>1.52) | 2.11<br>(0.85 -<br>5.89) | 1,167<br>(13%) | 673<br>(20%) | 1.66<br>(1.50 -<br>1.85) | 1.05<br>(0.69 -<br>1.58) |
| **Alcohol**<br>**consumed in**<br>**last 30 days**<br>**(11,997)** | No | 481 | 11300 | 1 (ref) | 1 (ref) | 8577 | 3204 | 1 (ref) | 1 (ref) |
|  | Yes | 5<br>(1.0%) | 211<br>(1.8%) | 1.80<br>(0.82 -<br>5.07) | 0.68<br>(0.11 -<br>13.5) | 132<br>(1.5%) | 84<br>(2.6%) | 1.70<br>(1.29 -<br>2.24) | 1.82<br>(0.68 -<br>4.79) |
| **Salt**<br>**consumption**<br>**(11,997)** | ≤ 5 grams | NA |  |  |  | 440<br>(5.1%) | 46<br>(1.4%) | 1 (ref) | 1 (ref) |
|  | > 5 grams |  |  |  |  | 8,269<br>(95%) | 3,242<br>(99%) | 3.75<br>(2.79 -<br>5.16) | **5.12**<br>**(2.30 -**<br>**13.7)** |
| **Sugar**<br>**consumption**<br>**(11,997)** | ≤ 50 grams | 440<br>(91%) | 8,269<br>(72%) | 1 (ref) | 1 (ref) | NA |  |  |  |
|  | > 50 grams | 46<br>(9.5%) | 3,242<br>(28%) | 3.75<br>(2.79 -<br>5.16) | **4.94**<br>**(2.22 -**<br>**13.2)** |  |  |  |  |
| **Fruit and**<br>**vegetable** | ≥ 5 servings<br>per day | 0 (0%) | 6<br>(<0.1<br>%) | 1 (ref) |  | 3<br>(<0.1%<br>) | 3<br>(<0.1<br>%) | 1 (ref) |  |
| ***servings per day (11,986)*** | < 5 servings per day | 484<br>(100%) | 11,496<br>(100%) | 0.00 |  | 8,698<br>(100%) | 3,282<br>(100%) | 0.38<br>(0.07 - 2.04) |  |
| ***Adequate physical activity (11,997)*** | No | 369 | 7736 | 1 (ref) | 1 (ref) | 6067 | 2038 | 1 (ref) | 1 (ref) |
|  | Yes | 117<br>(24%) | 3,775<br>(33%) | 0.65<br>(0.52 - 0.80) | 1.14<br>(0.62 - 2.05) | 2,642<br>(30%) | 1,250<br>(38%) | 0.71<br>(0.65 - 0.77) | 0.78<br>(0.58 - 1.05) |
| ***Stress (11,997)*** | Low stress | 121<br>(25%) | 2,459<br>(21%) | 1 (ref) | 1 (ref) | 2,032<br>(23%) | 548<br>(17%) | 1 (ref) | 1 (ref) |
|  | Moderate stress | 329<br>(68%) | 8,545<br>(74%) | 1.28<br>(1.03 - 1.58) | **1.97**<br>**(1.05 - 3.66)** | 6,322<br>(73%) | 2,552<br>(78%) | 1.50<br>(1.35 - 1.66) | **1.42**<br>**(1.01 - 2.00)** |
|  | High perceived stress | 36<br>(7.4%) | 507<br>(4.4%) | 0.69<br>(0.48 - 1.03) | 0.70<br>(0.27 - 1.94) | 355<br>(4.1%) | 188<br>(5.7%) | 1.96<br>(1.61 - 2.40) | **1.86**<br>**(1.04 - 3.32)** |
| ***Hypertension [19] (11,995)*** | Normal | 257<br>(53%) | 7,537<br>(65%) | 1 (ref) | 1 (ref) | 5,739<br>(66%) | 2,055<br>(63%) | 1 (ref) | 1 (ref) |
|  | Elevated | 39<br>(8.0%) | 986<br>(8.6%) | 0.86<br>(0.62 -<br>1.23) | **0.39**<br>**(0.18 -**<br>**0.91)** | 681<br>(7.8%) | 344<br>(10%) | 1.41<br>(1.23 -<br>1.62) | 0.95<br>(0.59 -<br>1.50) |
|  | HTN1 | 183<br>(38%) | 2,719<br>(24%) | 0.51<br>(0.42 -<br>0.62) | 0.56<br>(0.30 -<br>1.07) | 2,094<br>(24%) | 808<br>(25%) | 1.08<br>(0.98 -<br>1.19) | 1.17<br>(0.86 -<br>1.61) |
|  | HTN2 | 7<br>(1.4%) | 267<br>(2.3%) | 1.30<br>(0.66 -<br>3.07) | 0.63<br>(0.16 -<br>4.30) | 193<br>(2.2%) | 81<br>(2.5%) | 1.17<br>(0.90 -<br>1.52) | 0.94<br>(0.41 -<br>1.99) |
| **Depression**<br>**(11,997)** | No | 296 | 8403 | 1 (ref) | 1 (ref) | 6379 | 2320 | 1 (ref) | 1 (ref) |
|  | Yes | 190<br>(39%) | 3,108<br>(27%) | 0.58<br>(0.48 -<br>0.70) | 0.51<br>(0.26 -<br>1.02) | 2,330<br>(27%) | 968<br>(29%) | 1.14<br>(1.05 -<br>1.25) | **2.08**<br>**(1.48 -**<br>**2.92)** |
| **Anxiety**<br>**(11,997)** | No | 354 | 9726 | 1 (ref) | 1 (ref) | 7218 | 2862 | 1 (ref) | 1 (ref) |
|  | Yes | 132<br>(27%) | 1,785<br>(16%) | 0.49<br>(0.40 -<br>0.61) | 1.62<br>(0.67 -<br>4.43) | 1,491<br>(17%) | 426<br>(13%) | 0.72<br>(0.64 -<br>0.81) | 0.96<br>(0.63 -<br>1.46) |
| **BMI categories** <sup>[18]</sup><br>**(11,798)** | Normal | 269<br>(57%) | 6,508<br>(57%) | 1 (ref) | 1 (ref) | 4,940<br>(58%) | 1,837<br>(57%) | 1 (ref) | 1 (ref) |
|  | Sev Underweight | 40<br>(8.4%) | 857<br>(7.6%) | 0.89<br>(0.64 - 1.26) | 2.10<br>(0.60 - 13.3) | 675<br>(7.9%) | 222<br>(6.8%) | 0.88<br>(0.75 - 1.04) | 1.28<br>(0.77 - 2.11) |
|  | Underweight | 95<br>(20%) | 2,176<br>(19%) | 0.95<br>(0.75 - 1.21) | 0.99<br>(0.52 - 1.98) | 1,646<br>(19%) | 625<br>(19%) | 1.02<br>(0.92 - 1.14) | 0.83<br>(0.58 - 1.17) |
|  | Overweight | 33<br>(7.0%) | 919<br>(8.1%) | 1.15<br>(0.81 - 1.69) | 2.14<br>(0.78 - 7.67) | 665<br>(7.8%) | 287<br>(8.9%) | 1.16<br>(1.00 - 1.34) | 1.26<br>(0.79 - 1.99) |
|  | Obesity | 37<br>(7.8%) | 864<br>(7.6%) | 0.97<br>(0.69 - 1.39) | 1.68<br>(0.68 - 4.82) | 631<br>(7.4%) | 270<br>(8.3%) | 1.15<br>(0.99 - 1.34) | 1.08<br>(0.68 - 1.70) |
| **Diabetes**<br>**(1,599)** | Normal | 64<br>(85%) | 1,440<br>(94%) | 1 (ref) | 1 (ref) | 1,086<br>(94%) | 418<br>(93%) | 1 (ref) | 1 (ref) |
|  | Pre-diabetic | 9<br>(12%) | 70<br>(4.6%) | 0.35<br>(0.17 -<br>0.77) | **0.30**<br>**(0.13 -**<br>**0.71)** | 51<br>(4.4%) | 28<br>(6.2%) | 1.43<br>(0.88 -<br>2.27) | 1.50<br>(0.88 -<br>2.55) |
|  | Diabetic | 2<br>(2.7%) | 14<br>(0.9%) | 0.31<br>(0.08 -<br>2.01) | 0.58<br>(0.09 -<br>11.5) | 13<br>(1.1%) | 3<br>(0.7%) | 0.60<br>(0.14 -<br>1.87) | 0.51<br>(0.11 -<br>1.84) |
| No. Obs. |  | 1,291 |  |  |  | 1,291 |  |  |  |
|  |  | The model for stress shows odds adjusted for age category, sex, caste, wealth index, smokeless tobacco consumption, alcohol in past 30 days, sugar consumption (grams per person per day), fruit and vegetable consumption, stress, BMI categories, hypertension, diabetes, depression and anxiety |  |  |  | The model for stress shows odds adjusted for age category, sex, caste, wealth index, smokeless tobacco consumption, alcohol in past 30 days, sugar consumption (grams per person per day), fruit and vegetable consumption, stress, BMI categories, hypertension, diabetes, depression and anxiety |  |  |  |

OR = Odds Ratio, aOR= adjusted Odds Ratio CI = Confidence Interval, OBC = Other backward class, SC = Scheduled caste, ST = Scheduled tribe, VJ/NT/Others = Vimukt jati/Nomadic Tribe/Others, BMI = Body Mass Index, HTN = Hypertension, \*Indicates significant Odds ratios

A best fitting model was created for understanding predictors for sugar consumption which included age categories, gender, caste, marital status, education, wealth index, smokeless tobacco, alcohol and salt consumption, fruit and vegetable servings, physical activity, stress, hypertension, depression, anxiety, TG, LDL, HDL, BMI categories, diabetes status. Males had around 46% significantly higher odds of having excess sugar consumption compared to females. Those from SC and ST caste had significantly lower odds of 0.33 and 0.40 respectively of consuming excess amounts of sugar than those coming from Open caste. Those who were never married had 1.68 times more significant odds of consuming excess sugar than currently married respondents. For those who consumed excess salt per person per day, the odds of consuming excess sugar increased significantly by 5.12 times. Respondents who reported moderate and high levels of stress had 1.42 and 1.86 times higher odds respectively of consuming sugar in excess amounts than those who had low stress whereas those who reported depression had 2.08 times the odds for the same. The final model presented here had a Nagelkerke’s R^2^ value of 0.132 (table 6).

## Discussion

This paper discusses the baseline assessment of the study population with regards to the common modifiable risk factors of NCDs and the predictors of these risk factors in the age group of 10–30 years old individuals, which was carried out using the WHO STEPS tool. The characteristics such as occupation, education and marital status were non-comparable mainly due to age categories i.e. 10-20 and 20-30 years. The study identified the prevalence of some common behavioral and biochemical risk factors in the study area, which includes overall prevalence of the excess salt intake (95.9%), prevalence of smokeless tobacco users (15.3%) and smokers (0.3%), prevalence of alcohol consumers (1.8%), prevalence of inadequate physical activity (32.4%), prevalence of eating less than recommended servings of fruits and vegetables (99.99%), prevalence of obesity based on BMI (7.6%), prevalence of moderate (74%) and high (4.5%) perceived stress, prevalence of anxiety (16%) and depression (27.5).

Mean (SD) salt intake was 8.7 (3.1) grams per person per day in both the groups which is higher than the recommended salt intake by WHO [21] and it was comparable with the results from a multicentric study conducted in 2017 in India [22]. The average prevalence of smokeless tobacco (SLT) users was high (15.3%) as compared to smokers (0.3%). The prevalence of smokeless tobacco users was higher in the 20-30 age group i.e. 26.9% in rural areas which was also found in the Global Adult Tobacco Survey-2 published in 2020 [23]. This shows the need for early intervention in young age groups for better health outcomes in later life. The average prevalence of adequately physically active young individuals from both the age categories was very less i.e. approximately one third of the total, which was in sync with the studies conducted in the different parts of the world [24, 25]. The average number of fruits and vegetable servings per day is far less (1.5) than the expected minimum servings i.e. 5 servings for leading a healthy life as per WHO guidelines of healthy diet [26]. The average sugar consumption was found to be 44.7 grams per person per day which was less than the recommended limits by WHO i.e. 50 grams per day, but for additional health benefits, the sugar intake should be less than 5% of the total which is less than 25 grams per person per day (14), which indicates space for improvement in the daily eating habits.

The study showed that the age group 20-30 years had significantly higher risk of developing stress as compared to 10-20 years age group, whereas a study conducted in Gujarat, India on youth showed there is no significant relationship between age groups and stress in youth [27]. Male participants were having significantly lesser odds for developing stress as compared to female youth participants in the study which was also supported by a study conducted in Madhya Pradesh, India [28]. The odds of developing stress among OBC and Vimukt jati (VJ) was lower as compared to other castes. Though we did not find any article talking about the stress in VJ category people as compared to other castes, we did find articles about higher stress among scheduled caste people and OBC as compared to other higher Hindu castes which was opposite to what current study showed regarding OBCs[29, 30]. The study shows the association between education and stress in youth. As compared to no or primary education, secondary or higher education levels have higher chances of developing stress, which was also shown in various studies conducted in India [27, 31, 32]. The study also suggests that smokeless tobacco (SLT) consumers had higher odds of developing stress, which was aligned with a study conducted in India [33]. The study also suggested significantly higher odds of stress in participants with depression as compared with those who were not having depression which was also aligned with findings of a study conducted with school going adolescents in India [34]. But at the same time, study shows with decrease in anxiety levels, odds of having stress were increased, which contradicts the results of above-mentioned study [34].

The study shows a higher number of SLT consumers were males as compared to females. Similar results were also seen in the study from 2023 using data from Longitudinal Ageing Study in India (LASI) wave 1 and a report on tobacco control in India 2022 [35, 36]. Among students aged 13 to 15, 8.4% acknowledged currently using tobacco, as per the Global Youth Tobacco Survey 2 (GYTS), whereas The Global Adult Tobacco Survey-2 (GATS-2) found that tobacco use was more common as people aged, and this was true for both smoking and SLT items that contained tobacco, which aligns with the findings of the current study [36]. The occupation and its type showed a significant role in increased consumption of SLT. The unemployed, homemakers, students and service people have significantly lower odds of SLT consumption as compared to laborious work doing individuals. This finding also aligns with the findings from the LASI data study [35]. Additionally, compared to those with less education than graduation, the study indicates that those who have graduated and above had decreased odds of consuming SLT, which aligns with the findings from the LASI study [35]. However, as this study involves teenagers who were yet to complete their education, a large number of them might still be enrolled in school. Thus, it’s possible that bias might have been introduced here. The study shows significant inverse association between the SLT consumption and wealth index of the population which also coincides with the study conducted in India [37, 38]. The number of alcohol consumers using SLT was also more as compared to non-alcohol consumers, which also aligns with the findings from a study on National Sample Survey [39] while another study suggests the existence of spatial heterogeneity across different states and districts of India [40]. The study shows higher odds for consuming SLT with higher sugar consumption per day as well as moderate stress levels, but we could not find any previous study which can show relation between the respective variables. The current study also suggests the significant inverse association with the adequate physical activity and SLT consumption which also aligns with the findings from a study conducted in India [38]. The odds of having lower BMI (Severe underweight) are high and getting overweight are low in people consuming smokeless tobacco as compared to people with normal BMI which has also shown from the results of various studies conducted in India [41, 42].

The study shows statistically significant differences in physical activity status based on age category and gender. The study shows the level of inadequate physical activity decreases with increase in age group, while a study in Brazil shows that in adults, as the age increases the level of physical activity decreases [43]. Males were seen to be more physically active than females, which also aligns with findings from different studies conducted in India [25, 44, 45]. The study shows that the single people are having higher odds in engaging in inadequate physical activity as compared with the married and divorced participants, which contradicts the findings of a study conducted in Brazil [43]. The study suggests the participants with secondary education tend to have higher physical activity odds as compared to other levels of education, which is contradicting the findings from a study conducted in Spain [47]. A study also highlights the inverse relationship between physical activity and SLT consumption, which coincides with the findings from the study conducted in India [38]. It was found that alcohol consumers are less likely to have inadequate physical activity as compared to those without alcohol consumption, which coincides with the findings from a Brazilian study [47]. The study shows that adolescents with moderate levels of stress are more predisposed to indulge in physical activity. Physical activity was found to be more inadequate in those having anxiety, whereas the study conducted with a group of female students in 2020 showed no effect of high physical activity on stress levels but aligned with this study results regarding the correlation between physical activity and anxiety [48]. The study suggests higher odds of having hypertension with higher inadequate physical activity, which aligns with the findings from a review [49]. The study shows higher odds of having inadequate physical activity with severely underweight people which lower odds of inadequate physical activity with obese people, which is contradicting with the findings from the study conducted in the UK [50]. Even though the study shows that those who consume more salt and sugar are having a negative likelihood of inadequate physical activity, any direct association of the same was not found.

The study shows statistically significant differences in sugar consumption in case of gender i.e. Males have significantly higher odds of consuming appropriate amounts of sugar as compared to females, but a study conducted in a city of Karnataka state with school going adolescents showed no significant difference based on gender [51]. The difference in the two studies might be due to the study setting being different i.e. rural vs urban. The level of sugar intake was significantly more in unmarried as compared with married participants, which was also the findings of the study conducted in an English longitudinal study of aging [52]. The study also indicates significantly higher odds of appropriate consumption of salt in unmarried participants as compared with married, whereas the study conducted in Delhi shows no significant relationship between the salt consumption and marital status of the participant women [53], but a study conducted in Saudi Arabia showed contradictory results as compared to the results of our study [54]. The study signifies association of salt consumption with wealth index as the second quintile of the wealth index has significantly higher odds of appropriate salt intake as compared with lowest, while a study conducted on south Indian population showed higher the income, higher would be the salt consumption [55]. The study showed elevated blood pressure (BP) significantly decreases the odds of having appropriate salt consumption as compared to normal BP. The participants with moderate stress level were having higher chances of consuming appropriate amounts of salt as compared to low stress people, but the review published in 2010 showed that acute stress has no effect on salt consumption, however, chronic stress may have effect on salt consumption [56]. The SC/ST caste showed lesser odds for the consumption of appropriate amounts of sugar. We were unable to find this relation between these two variables in the literature. The study shows higher odds for consuming appropriate amounts of salt in high SLT consuming people, but we could not find any previous study which can show relation between the two variables.

## Strengths and limitations

A large sample was surveyed through simple random sampling which reduces potential risk of selection bias. Our study has used the WHO STEPS questionnaire which is a standardized tool which makes the results of this study comparable with other international studies. Thus, the data analysis was performed as per the standard cutoffs and guidelines available for analysis like the analysis guide was used for analyzing data on physical activity. Very few such studies are available from India which have been conducted on such a large sample in this age group. We weren’t able to find Indian reference values for biochemical tests in adolescent age groups and thus have used American cut-offs for a few variables. The cut-offs suggested by our study can be tested on a larger sample size for correctness and standardization for Indian use.

| Sr. No. | What is known | This study adds to the existing body of knowledge |
| --- | --- | --- |
| 1. | Common risk factors for NCDs | Risk factors for risk factors of NCDs |
| 2. | Lifestyle habits develop during the early years of life and influenced during adolescence | Strength of association between the various risk factors |
| 3. | Prevalence of NCDs in adults and little literature around prevalence of risk factors and NCDs in adolescents | Prevalence of risk factors as well as hypertension and diabetes in this age group on such a large sample |
| 4. |  | Our study includes not only the behavioural risk factors but also the biochemical risk factors in a sub group of 1600 study participants. |

## Conclusion

The prevalence of risk factors for non-communicable diseases (NCDs) is notably high among adolescents, underscoring the importance of frequent screening through comprehensive surveys. Such proactive measures can enable timely interventions aimed at reducing the incidence of NCDs. While current policies predominantly address tobacco consumption through smoking regulations, there is a pressing need for more stringent laws concerning the use of chewable tobacco. Strengthening the screening of adolescents for various NCDs and their associated risk factors into existing programs, such as the Rashtriya Kishor Swasthya Karyakram (RKSK), Rashtriya Bal Swasthya Karyakram (RBSK), National Programme for Non-communicable diseases (NP-NCD), the National Family Health Survey (NFHS) and Global Youth Tobacco Survey (GYTS) can ensure that this issue is recognized as a significant public health concern. By doing so, we can take timely, policy-based actions to address and mitigate the impact of NCDs among the youth.

## Data Availability

The data that support the findings of this study are available from the corresponding author upon reasonable request.

## Acknowledgements

We would like to thank the CEO (Wardha), district authorities from departments of education, PRI and MSRLM for their cooperation and support. Also, we acknowledge the support of all the community members including VCaN club members as well as all villagers who enthusiastically participated in all the activities during the project, our surveyors and data enumerators.

## Declaration

None of the authors have any conflict of interest.

## Trial Registration details

Clinical Trials Registration India CTRI/2020/10/028700; https://ctri.nic.in/Clinicaltrials/showallp.php?mid1=47597&EncHid=&userName=V-CaN International Registered Report Identifier (IRRID): DERR1-10.2196/42450

